# “Real-time county-aggregated wastewater-based estimates for SARS-CoV-2 effective reproduction numbers”

**DOI:** 10.1101/2024.05.02.24306456

**Authors:** Sindhu Ravuri, Elisabeth Burnor, Isobel Routledge, Natalie Linton, Mugdha Thakur, Alexandria Boehm, Marlene Wolfe, Heather N. Bischel, Colleen C. Naughton, Alexander T. Yu, Lauren A. White, Tomás M. León

## Abstract

**Background:** The effective reproduction number (*R_e_*) serves as a metric of population-wide, time-varying disease spread. During the COVID-19 pandemic, *R_e_* was primarily estimated from clinical surveillance data streams (*R_cc_*), which have varied in quality and representativeness due to changes in testing volume, test-seeking behavior, and resource constraints. Deriving *R_e_* from alternative data sources such as wastewater could inform future public health responses.

**Objectives:** We estimated county-aggregated, sewershed-restricted wastewater-based SARS-CoV-2 *R_e_* (*R_ww_*) from May 1, 2022 to April 30, 2023 for five counties in California of varying population sizes, clinical testing rates, demographics, proportions surveilled by wastewater, and sampling frequencies to validate the reliability of *R_ww_* as a real-time disease surveillance metric.

**Methods:** We produced both instantaneous and cohort sewershed-restricted *R_e_* using smoothed and deconvolved wastewater concentrations. We then population-weighted and aggregated these sewershed-level estimates to arrive at county-level *R_e_*. Using mean absolute error (MAE), Spearman’s rank correlation (ρ), confusion matrix classification, and cross-correlation analyses, we compared the timing and trajectory of two *R_ww_* models to: (1) a publicly available, county-level ensemble of *R_cc_* estimates, and (2) a county-aggregated, sewershed-restricted *R_cc_*.

**Results:** Both *R_ww_* models demonstrated high concordance with traditional *R_cc_* estimates, as indicated by low mean absolute errors (MAE ≤ 0.09), significant positive Spearman correlation (Spearman ρ ≥ 0.66, p < 0.001), and high confusion matrix classification accuracy (≥ 0.81). The relative timings of *R_ww_*and *R_cc_* were less clear, with cross-correlation analyses suggesting strong associations for a wide range of temporal lags that varied by county and *R_ww_* model type.

**Discussion:** This *R_e_* estimation methodology provides a generalizable, robust, and operationalizable framework for estimating county-level *R_ww_*. Our results support the additional use of *R_ww_*as an epidemiological tool for surveillance. Based on this research, we produced publicly available *R_ww_* nowcasts for the California Communicable diseases Assessment Tool (https://calcat.covid19.ca.gov/cacovidmodels/).

## INTRODUCTION

The effective reproduction number (*R_e_*) of a disease represents the average number of secondary infections caused by a newly infectious individual within a population of both susceptible and immune hosts.^1^ An *R_e_* value greater than one suggests the number of new infections within a population will increase, while an *R_e_* value less than one indicates the number of new infections will decrease.^2^ In the context of SARS-CoV-2, *R_e_* commonly serves as a metric of population-wide disease spread under conditions of time-varying vaccination rates, immunity viral variant evolution, health protective behavior modifications, and other response measures.^2^ Importantly, monitoring *R_e_* can inform public health policy decisions (e.g., travel restrictions, school closures, mask requirements).^3–6^

In the first years of the COVID-19 pandemic, *R_e_* was estimated from clinical surveillance data streams such as confirmed case counts (i.e., “case-based” *R_e_*). However, the quality and representativeness of clinical data are not always consistent through time and space. Ongoing changes in testing capacity, access, eligibility, test-seeking behavior and reporting may bias case-based *R_e_* estimates; for example, a simulation study mimicking alterations to testing practices found that increasing or decreasing the proportion of detected cases over- or under-estimated *R_e_*, respectively.^7^ Delays and administrative noise in case reporting (e.g., lagged processing speed for weekend data) are also common issues for traditional *R_e_* estimation tools.^8^ Deriving *R_e_* from alternative data sources will be an important means of informing response to the future public health impact of COVID-19 and bolstering nowcasting efforts.

The COVID-19 pandemic highlighted wastewater surveillance as a timely and accurate monitoring tool reflecting community-wide disease transmission.^9,10^ Wastewater surveillance measures the amount of SARS-CoV-2 viral RNA shed into wastewater by infected individuals. It circumvents limitations of traditional clinical SARS-CoV-2 surveillance in important ways. First, it captures both asymptomatic and symptomatic infections.^11^ Second, wastewater surveillance is not as impacted by heterogeneous, time- and space-varying testing-related factors.^12–16^ Third, wastewater surveillance data is usually available within days, overcoming case reporting delays.^12–16^ For these reasons, using wastewater surveillance could improve the accuracy and reliability of *R_e_* estimates while also compensating for data delays from traditional sources. In addition, given that wastewater surveillance measures viral RNA concentrations at the level of a single wastewater treatment plant’s catchment area (sewershed), it could be used to estimate real-time *R_e_* in finite geographies that may lack robust case reporting.

Existing methods of wastewater-based *R_e_* (denoted as *R_ww_* in this study, following notation by Huisman et al.^17^) estimation include SEIR-like compartmental models,^18^ branching process-inspired models,^17,19,20^ and artificial neural networks.^21^ Despite using different underlying mathematical approaches, these methods have consistently revealed high concordance between *R_ww_* and conventional case-based *R_e_* estimates (denoted as *R_cc_* in this study, following notation by Huisman et al.^17^). In some circumstances, *R_ww_*even highlights outbreak dynamics not captured by *R_cc_* alone; for example, Nadeau et al.^20^ demonstrated that, compared to *R_cc_*, *R_ww_* better reflected the impact of COVID-19 interventions on influenza transmission. Notably, these studies have typically examined *R_ww_* at the scale of an individual sewershed. However, a single county may contain multiple treatment plants representing distinct communities, and sewersheds do not typically map to municipal boundaries. Moreover, public health policies are often enacted on a county or regional level. Deriving county-level *R_ww_* estimates that integrate data from multiple sewersheds would therefore provide a more comprehensive view of community transmission dynamics at a geographic scale relevant for public health policymaking.

In this study, we provide proof-of-concept estimation of county-aggregated, sewershed-restricted *R_ww_* for the state of California from May 1, 2022 to April 30, 2023. We compare the timing and trajectory of two *R_ww_* models to: (1) a publicly available, county-level ensemble of *R_cc_* estimates, and (2) a county-aggregated, sewershed-restricted *R_cc_* for SARS-CoV-2 (though it is important to note that *R_ww_* and *R_cc_* are both measuring an unobservable quantity, *R_e_*, the “true” underlying parameter). To evaluate generalizability of our estimation procedure across distinct populations, we present results for five counties with varying sizes, clinical testing rates, demographics, proportions surveilled by wastewater, and sampling frequencies.

## METHODS

### R_e_ Estimation Overall Approach

To produce *R_ww_* estimates, we relied on the *R_e_* derivation frameworks developed by Cori et al.^1^ and Wallinga & Teunis^22^ (hereafter denoted as instantaneous or cohort *R_e_*, respectively). These two modeling approaches, traditionally applied on case data, have been well characterized and broadly used in public health for real-time *R_cc_*estimation.^8^

We compared the timing and trajectory of these two *R_ww_*models to the following: (1) county-level *R_cc_* ensembles sourced from the California Communicable diseases Assessment Tool, CalCAT (https://calcat.covid19.ca.gov/cacovidmodels/) (hereafter denoted as the CalCAT ensemble); and (2) *R_cc_* estimates produced by applying Cori et al.^1^ and Wallinga & Teunis^22^ to sewershed-restricted case data (hereafter denoted as sewershed-restricted *R_cc_*). Comparing *R_ww_* results to the CalCAT ensemble allows us to validate the robustness and reliability of *R_ww_* as a real-time disease surveillance metric for public health. Additionally, comparing *R_ww_* results to sewershed-restricted *R_cc_* offers two potential advantages. First, sewershed-restricted *R_cc_* represents the same populations within a given county as *R_ww_*, while the CalCAT ensemble relies on county-wide data sources. Second, sewershed-restricted *R_cc_* can be indexed by time of infection. In contrast, the CalCAT ensemble integrates different models with varying *R_e_* estimation procedures, meaning its estimates cannot be easily indexed by time of infection. Therefore, comparing *R_ww_* and sewershed-restricted *R_cc_* allows us to contextualize the former’s timing with respect to conventional case-based approaches.

We first conducted our *R_ww_* estimation process using wastewater data. We subsequently repeated the estimation procedure on sewershed-restricted case data to produce *R_cc_* (Figure 2). County-level CalCAT ensemble estimates are publicly available and were not further processed. Our analysis period spanned one year (May 1, 2022 to April 30, 2023).

### R_e_ Estimation Process

#### STEP 0: Raw Data Sources

#### Wastewater Sampling

Wastewater samples included in the study were collected during the analysis period using two different methods of sample processing, as described below (Table 1). Samples were collected from 14 sewersheds in five California counties at a frequency of three to seven times per week (Table 1). Samples from eight sites (San Francisco Southeast, San Francisco Oceanside, Sacramento, Palo Alto, San Jose, Sunnyvale, Gilroy/Morgan Hill, and Davis) were processed according to Method 1. Samples from six sites (Modesto, Merced, Turlock, Esparto, Winters, and Woodland) were processed according to Method 2 from May 1, 2022 to November 30, 2022 and according to Method 1 from December 1, 2022 until April 30, 2023 (Tables 1 and S1). Method 2 was designed to closely follow the protocol of Method 1, with sample processing and laboratory analyses performed at a different laboratory.

**Table 1.**
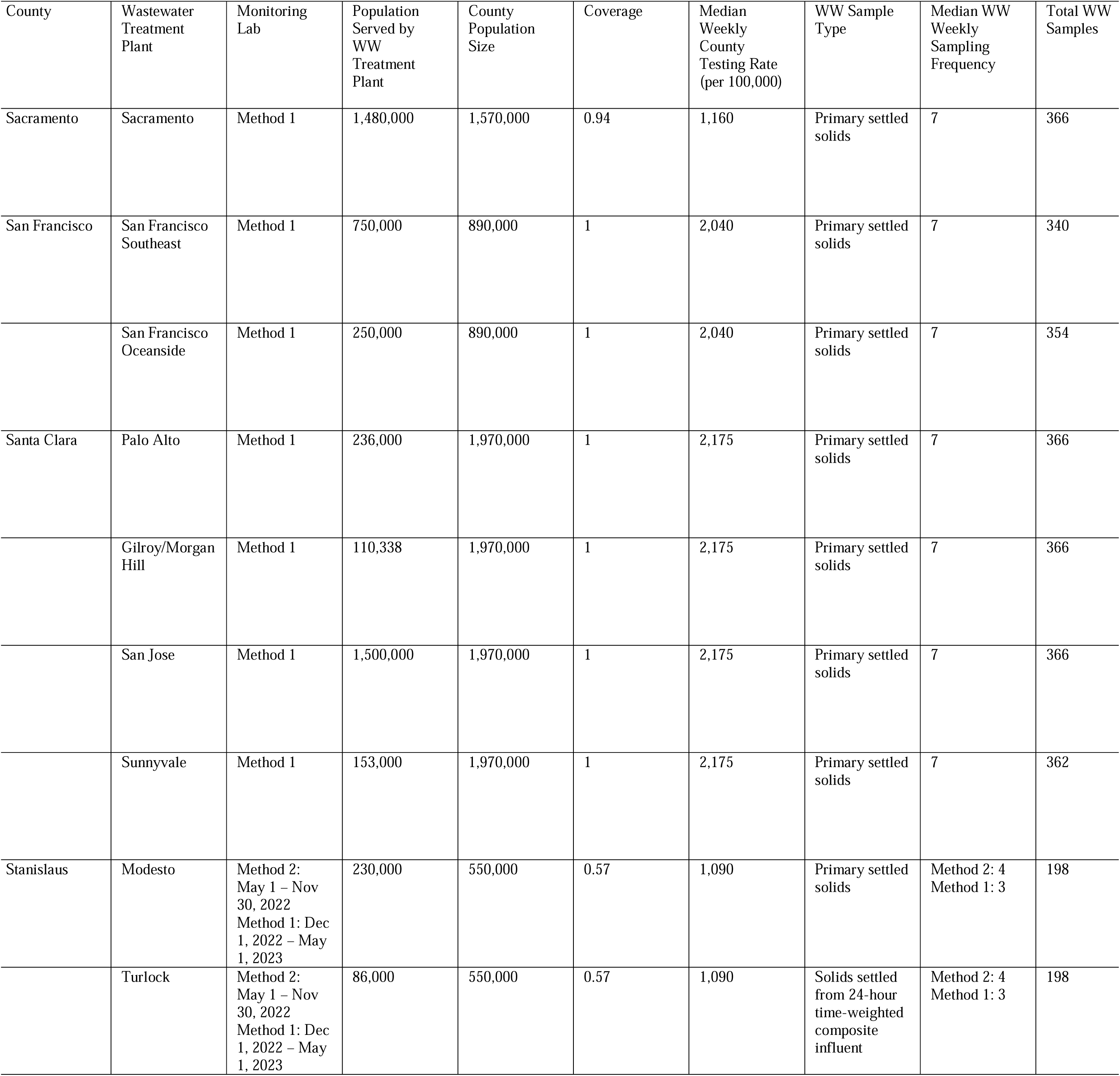
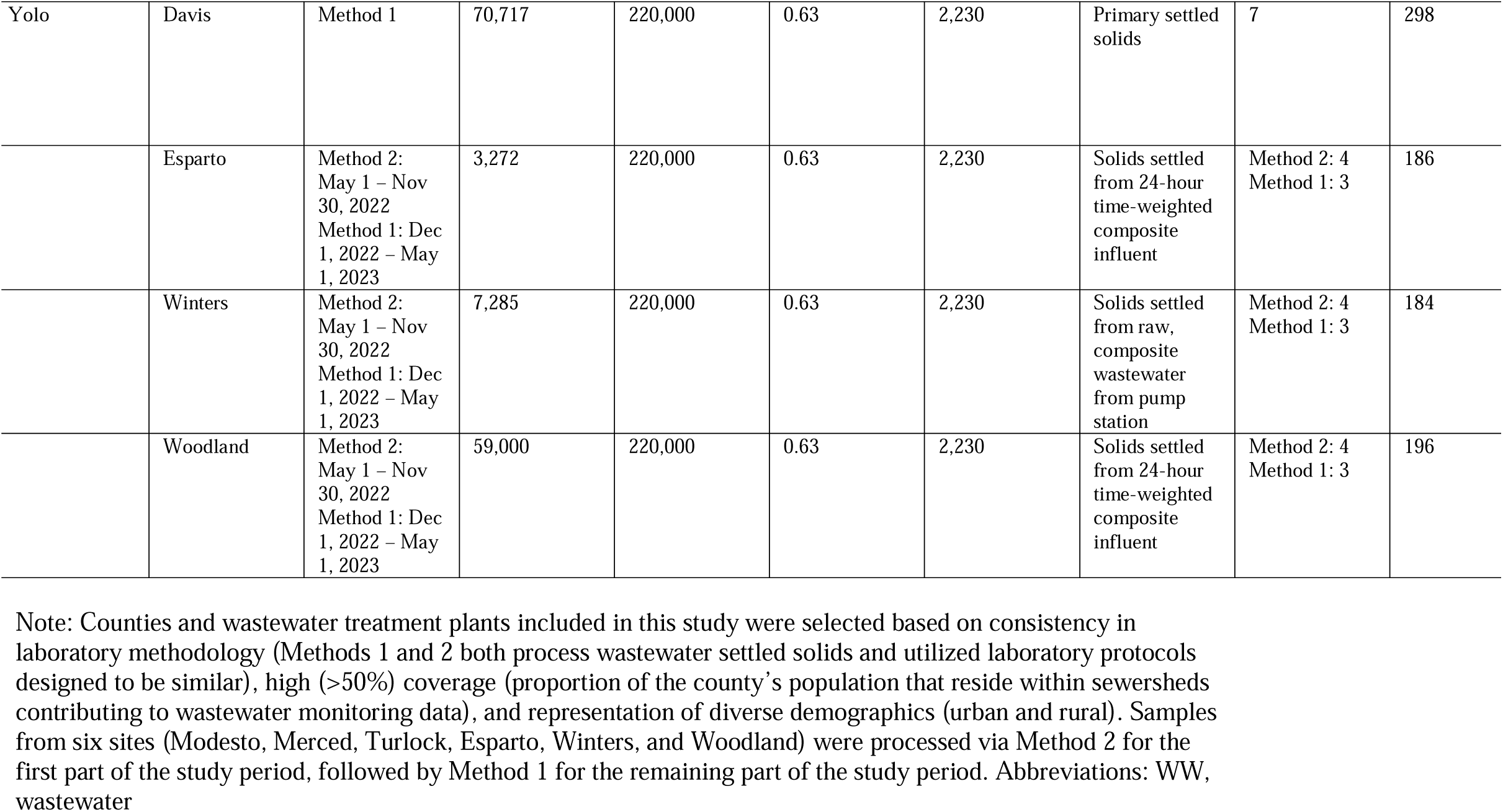
Population and sampling characteristics of counties and wastewater treatment plants included in the analyses.

##### Method 1, Sample Processing

Protocols developed for Method 1 were designed and reported according to the Environmental Microbiology Minimum Information (EMMI) guidelines, as described in Borchardt et al.^23^ For samples processed via Method 1, 50 milliliters of wastewater settled solids were collected three to seven times per week from the primary clarifier or from influent using an Imhoff cone,^24,25^ transferred at 4°C and processed immediately upon receipt at the lab. The methods used to measure the N gene via digital droplet RT-PCR, including thorough descriptions of the extraction and PCR negative and positive controls, process control recovery, QA/QC elements, thresholding methods, and relevant EMMI guideline reporting, have been described in detail in the Supplemental Materials and in a previously published data descriptor.^26^

Briefly, wastewater settled solids were dewatered with centrifugation and added to DNA/RNA Shield (Zymo Research Corporation, Irvine, CA) at a concentration of 75 mg/ml to minimize inhibition. Nucleic acids were extracted and quantified by a digital droplet RT-PCR assay (dd-RT-PCR) targeting the N gene of SARS-CoV-2.^26^ More detailed methods can be found in the Supplemental Materials.

##### Method 2, Sample Processing

For samples processed via Method 2, the collection procedure of wastewater solids varied by site (Table 1). For Modesto, grab samples of settled solids from a primary clarifier were collected (similar to Method 1). For Esparto, Turlock, and Woodland, composite samples of influent wastewater were collected, and for Winters, composite samples of raw wastewater were collected from a pump station (Table 1). For sites collecting liquid wastewater samples, solids were obtained by settling the samples in a glass beaker for a minimum of 30 minutes. These settled solids were then processed according to the same protocol described for primary settled solids.^27^ For all sites, samples were collected three to five times per week and transferred at 4°C to the laboratory. Settled solids were dewatered, diluted in DNA/RNA Shield (Zymo Research Corporation, Irvine, CA), extracted, and quantified by dd-RT-PCR. Minor differences in the processing of dewatered solids, extraction, and dd-RT-PCR assays between Methods 1 and 2 are described in Kadonsky et al.^27^ A comparative interlaboratory analysis of wastewater samples collected from sewersheds in Davis city of Yolo county suggests that data obtained using Methods 1 and 2 are comparable.^27^

In this analysis, we used raw, unadjusted wastewater data in units of copies of SARS-CoV-2 RNA per gram of wastewater solids.

#### County Selection

For this analysis, we estimated *R_ww_* for five California counties: Sacramento, San Francisco, Santa Clara, Stanislaus, and Yolo. We selected counties using similar laboratory methodologies that have one or more site(s) with long wastewater sampling histories (multiple years of available data). Inter-method comparisons of wastewater-derived SARS-CoV-2 concentrations can be highly complex, as quantification is dependent on several unique features of a processing pipeline (e.g., sample type (liquid vs. solids), extraction method and efficiency, assay target selection, quantity of extraction replicates and PCR replicates, laboratory equipment, etc.^28^ Although six of our sites underwent laboratory changes and all sites underwent minor changes in methodology during the study period, we sought to avoid some of the complications of inter-method data comparability by including only sites processing settled solids (either grab samples from a primary clarifier or solids settled out from a composite wastewater sample) and only sites using the same assay target for SARS-CoV-2 (see Supplemental Methods).

Wastewater monitoring tends to be more robust in urban locations for reasons such as larger population coverage, proximity to laboratories, and greater plant staffing and resources, resulting in inequitable coverage for rural communities.^29^ Therefore, we included counties from both urban (San Francisco, Palo Alto, and Sacramento) and rural areas (Stanislaus and Yolo) (Figures 1 and S1).

**Figure 1.**
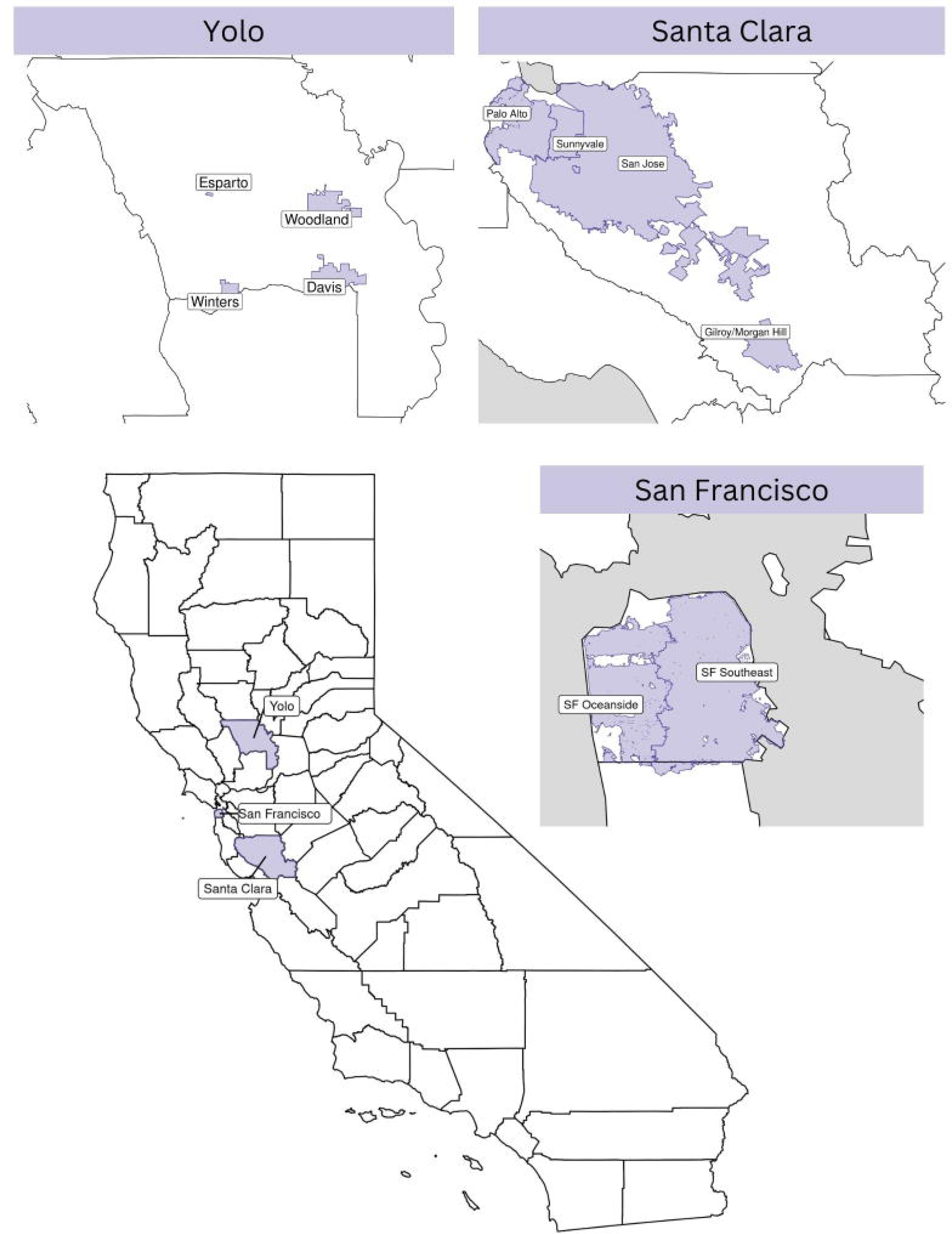
Map of sewersheds included in analysis. The geographical boundaries of sewersheds (a wastewater treatment plant’s catchment area) selected for the present study are highlighted in blue. Each sewershed’s respective county is outlined in black. Three of the five studied counties (San Francisco, Santa Clara, and Yolo) are illustrated; maps of the remaining counties (Sacramento and Stanislaus) are included in the Supplemental Material.

**Figure 2.**
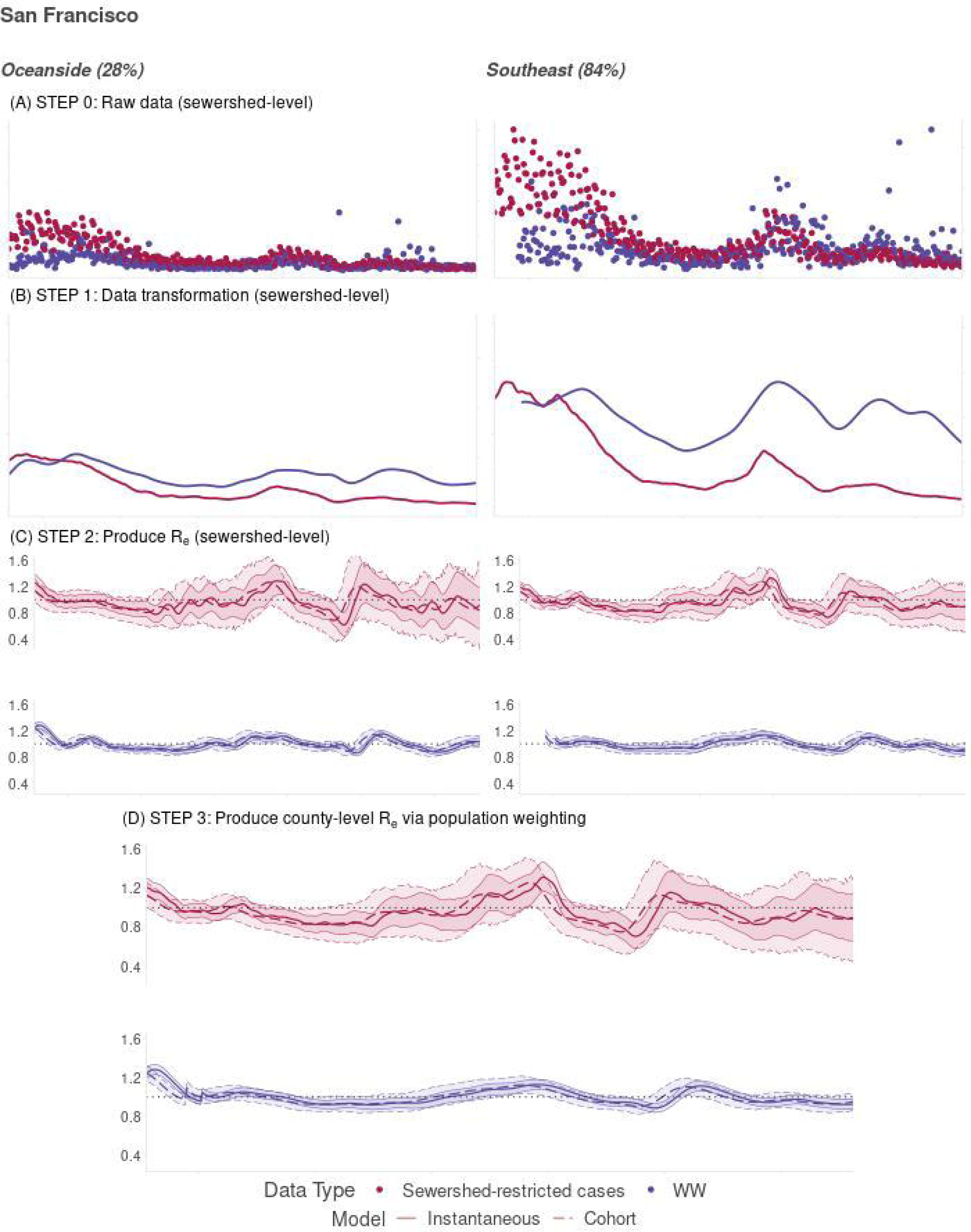
Stepwise process of county-aggregated, sewershed-restricted *R_e_* estimation for a single location. Using San Francisco as an example, we demonstrate our stepwise pipeline for estimating county-aggregated, sewershed-restricted *R_e_*. Each column represents a single wastewater treatment plant within the county. The left column corresponds to San Francisco Oceanside, which surveils 28% of the total county population; the right column corresponds to San Francisco Southeast, which surveils 84% of the total county population. Detailed axes are excluded for this generic, representation of the *R_e_* estimation process. (A) The pipeline begins with raw, sewershed-restricted time series data; in our study, input data includes case counts (pink) and wastewater viral concentrations (blue). (B) In Step 1, raw time series data are transformed: case counts are LOESS smoothed, while wastewater concentrations are spline smoothed and root transformed. Transformed data are subsequently deconvolved, shifting both time series’ backwards temporally such that observations are indexed by date of infection. (C) In Step 2, the methods of Cori et al.^1^ and Wallinga and Teunis^22^ are applied on the modified time series data streams to produce instantaneous and cohort sewershed-level *R_e_* estimates. (D) Finally, in Step 4, sewershed-level *R_e_* estimates are population-weighted and aggregated to yield county-level *R_e_* estimates. Note: WW, wastewater surveillance data; *R_e_ ,* effective reproduction number; Model, *R_e_* modeling approach (instantaneous or cohort).

We define “coverage” as the proportion of a county’s population that resides within sewersheds that are monitored with wastewater surveillance. Amongst California counties, coverage varies widely, ranging from as low as 10% to as high as 100%. *R_ww_* estimates produced for counties with low coverage may be subject to selection bias, as they only account for infections in small subpopulations of the county that are sampled via wastewater monitoring programs. Such *R_ww_* estimates may consequently follow a trajectory that does not accurately reflect county-wide infection and transmission dynamics. Due to this concern, we chose counties with at least 50% coverage.

#### Sewershed-Restricted Case Data

Sewershed shapefiles were generated in collaboration with each of the wastewater treatment plants selected for this analysis. PCR-confirmed COVID-19 case counts reported to the California Department of Public Health (CDPH) were linked with each sewershed using methods described previously.^30^ Cases were counted as a function of episode date (earliest of date received, date of diagnosis, date of symptom onset, date of death, or date of specimen collection). The California Health and Human Services (CHHS) Committee for the Protection of Human Subjects (CPHS) determined that use of this data is exempt from review under their criteria.

#### CalCAT Ensemble

As part of the COVID-19 Response, CDPH developed a *R_e_* nowcast ensemble publicly available on CalCAT (https://calcat.covid19.ca.gov/cacovidmodels/). This nowcast is a smoothed median ensemble of both internally generated and externally contributed models. Median ensembles tend to outperform individual models, as has been demonstrated during the COVID-19 pandemic.^31,32^ Smoothing ensures that the ensemble is robust to outliers, dampening noise in the median nowcast. The models are predominantly case-based, though some additionally or instead use test positivity, hospitalizations, intensive care unit census, and/or deaths as an input (Table S2).

The CalCAT ensemble is produced on county, region, and state levels. In this study, we strictly use county-level CalCAT ensemble estimates. The ensemble is retroactively updated for the dates through the present day, based on latest estimates from contributing models. The number and cadence of daily contributor models to the ensemble may vary across time and geographies.

The CalCAT ensemble continues to serve as a real-time disease dynamics monitoring tool informing state policymakers on the speed and strength of SARS-CoV-2 transmission and burden.

#### STEP 1: Data transformation

Input time series data streams for sewershed-level *R_e_*estimation (i.e., case counts and wastewater viral concentrations) were transformed using distinct approaches to minimize noisiness for each data stream (Figure 2). Case counts were LOESS smoothed using the R package estimateR. Raw wastewater concentrations were spline smoothed using the R package npreg.^33^ To fit sewershed-specific smoothing splines, we spaced knots by seven days over the course of the full analysis period. We selected optimal smoothing parameters unique to each sewershed using ordinary cross validation. Smoothed wastewater concentrations were subsequently square root-transformed. Root transformation produced *R_ww_* estimates that were more comparable in absolute terms to *R_cc_*, as raw wastewater concentrations (which can be as high as 10^6^ copies/gram) are often much greater in magnitude and variability than traditional case counts.

#### STEP 2: Produce sewershed-level *R_e_*

Both instantaneous and cohort *R_e_* models required a time series input of incidence data indexed by date of infection as well as an estimate of the generation time (i.e., the time interval between infections of an infector and their infectee(s)).^1,34^ Sewershed-restricted case counts were indexed by episode date, while wastewater concentrations were indexed by date of shedding into wastewater. We re-indexed these data streams to date of infection using deconvolution with input-specific delay distributions. For case data, we derived a delay distribution using the California COVID-19 case registry. As is typical of observational line list data, date of infection was unknown, and date of symptom onset was not available for many cases, in large part due to the presence of mild or asymptomatic infections and reporting practices. Since episode date in the line list was most representative of the date of nucleic acid amplification test (NAAT) result, we approximated episode date as the date of NAAT result. Next, we deconvolved from date of NAAT result to date of infection using two delay distributions: (1) an estimate of the incubation period, based on Aguila-Mejia et al.,^35^ and (2) the delay from symptom onset to NAAT result, based on the line list (Table 2). For wastewater, we used the infection to shedding distribution reported by Huisman et al. (Table 2).^17^ This distribution was optimized for San Jose, California, and reflects the profile of SARS-CoV-2 RNA viral shedding by an infected individual during the days post-infection. We used the R package estimateR to perform deconvolution through a variant of the Richardson-Lucy expectation-maximization algorithm.^36^

**Table 2.** Key distribution parameters used in sewershed-level *R_e_* estimation pipeline.

| Distribution | Purpose | Relevant $R_e$ | Distribution Type | Mean (SD) | Distribution Parameters | Source |
| --- | --- | --- | --- | --- | --- | --- |
| Generation time | Sewershed-restricted $R_e$ estimation from input time series | $R_{ww}$; sewershed-restricted $R_{cc}$ | Gamma | 6.84 (4.48) | Shape: 2.33<br>Scale: 2.93 | Manica et al.<sup>39</sup> |
| Infection to shedding | Deconvolution of wastewater concentrations | $R_{ww}$ | Gamma | 5 (0.5) | Shape: 100<br>Scale: 0.05 | Huisman et al.<sup>17</sup> |
| Incubation period | Deconvolution of sewershed-restricted case counts | sewershed-restricted $R_{cc}$ | Lognormal | 3.1 (2.6) | Meanlog: 0.87<br>Sdlog: 0.73 | Aguila-Mejia et al.<sup>35</sup> |
| Onset to NAAT result | Deconvolution of sewershed-restricted case counts | sewershed-restricted $R_{cc}$ | Lognormal | 3.35 (2.84) | Meanlog: 0.94<br>Sdlog: 0.73 | CA cases line list |

Note: Estimation of instantaneous and cohort sewershed-level $R_e$ requires a user-specified generation time distribution. Estimation also requires a time series input of incidence data indexed by date of infection. To index both raw sewershed-restricted case counts and wastewater viral concentrations by date of infection, we performed deconvolution. For cases, we deconvolved using two delay distributions. For wastewater, we deconvolved using a single delay distribution optimized for San Jose, California. Abbreviations: NAAT, nucleic acid amplification test, $R_e$ , effective reproduction number; $R_{ww}$ , sewershed-restricted, wastewater-based effective reproduction number; $R_{cc}$ , case-based effective reproduction number; SD, standard deviation.

We also used the estimateR package to produce *R_e_* values derived from the Cori et al.^1^ method (hereon referred to as instantaneous *R_ww_*). estimateR implements a wrapper around the Cori et al.^1^ approach and enables repeated *R_e_* estimation on a user-defined number of bootstrap samples built from the input data time series.^36^ The distribution of bootstrapped *R_e_* estimates is then used to produce 95% confidence intervals.^36^ We generated 1000 bootstrap samples for our study for each location estimate. To produce *R_e_* values derived from the Wallinga and Teunis method (hereon referred to as cohort *R_ww_*), we used the R package R0.^37^ We applied an adjustment for right-censorship to obtain accurate *R_e_*estimates at the end of the time series.^34,38^ With the R0 package, we ran 1000 multinomial simulations at each time step to compute 95% confidence intervals.^37^ Both the instantaneous and cohort models exhibit noise in initial *R_e_* estimates (i.e., extreme spikes). To account for this, we trimmed the first week of estimates for each sewershed-level *R_e_*. For both models, we utilized the generation time reported by Manica et al. (Table 2).^39^

#### STEP 3: Produce county-level *R_e_* via population weighting

Sewershed-restricted *R_ww_* and *R_cc_* estimates were weighted by population and subsequently aggregated to produce county-level *R_e_*. The aggregation approach is described in Equation [1], where *R_ww_,_i_* represents *R_ww_* of sewershed *i*, *Pop_i_* represents the population size serviced by sewershed *i*, and *n* represents the total number of sewersheds enrolled in wastewater monitoring within the relevant county. This aggregation approach ensures sewersheds corresponding to larger or smaller populations have proportional influence on and representation in the county-level *R_ww_* estimate.

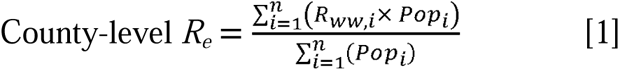

Sewershed population sizes should be considered approximate, as they are estimated by wastewater utilities using unique and non-standardized methodologies.

For Yolo county’s *R_cc_*, we modified our aggregation approach to accommodate low case counts in sewershed-restricted boundaries. The minimum number of cumulative infections prior to *R_e_* estimation recommended by Cori et al. is 12.^1,36^ In certain Yolo sites (Esparto and Winters), the minimum number of cumulative infections prior to *R_e_*estimation on May 1, 2022 (the analysis period start date) was below this threshold. Such low input case counts generated unreliable *R_cc_* estimates with extreme sensitivity to small shifts in case incidence. To address this, we combined case counts across all Yolo sites and input this single time series into our *R_e_* estimation procedure.

Only sewersheds with wastewater data available for a given date were included in the county-aggregated *R_e_* estimate for that date. While most sewersheds in our analysis sampled wastewater for the full study period, San Francisco Southeast began sampling on May 20, 2022. As a result, county-aggregated *R_ww_* for San Francisco only includes San Francisco Oceanside for the first 19 days of the study period.

We derived confidence intervals for county-level *R_ww_*and *R_cc_* as population-weighted ensembles of sewershed-level confidence intervals. For each date, we assumed that a sewershed’s *R_e_* (*R_ww_* or *R_cc_*) point estimate served as the mean of a normal (or Gaussian) distribution with 95% confidence intervals matching those estimated for each sewershed. Under this simplifying assumption, we then linearly interpolated the means and variances of sewershed-level normal distributions within a county to produce a combined normal distribution; confidence intervals of this resultant distribution were used for county-level *R_ww_* and *R_cc_*. This method of linear interpolation of Gaussian distributions has been shown to reduce the error coverage for ensemble estimates.^40,41^ Uncertainty estimation for the CalCAT ensemble was not feasible, as contributing models did not all report confidence intervals.

### Comparison of R_e_ trajectories

To evaluate *R_ww_* and *R_cc_* concordance in magnitude and direction, we calculated the mean absolute error (MAE) and Spearman’s rank correlation (ρ). For *R_ww_* versus sewershed-restricted *R_cc_*, identical *R_e_* models (instantaneous or cohort) were compared.

CalCAT ensemble values provide real-time, ongoing *R_cc_*estimates with relevance for statewide public health response, while sewershed-restricted *R_cc_* values provide comparative controls for our particular study. Additional analyses characterizing the directional concordance between *R_ww_* and the CalCAT ensemble would provide greater evidence of the former’s potential real-time public health utility. To that end, we produced multi-class confusion matrices relating *R_ww_*to the CalCAT ensemble. Based on magnitude, *R_e_* values were grouped into transmission strength categories for the state of California (<0.7, 0.7-0.9, 0.9-1.1, 1.1-1.3, >1.3, which represent a sharp decrease, decrease, stability, increase, and sharp increase in *R_e_*, respectively). When predicted *R_ww_* values and reference CalCAT ensemble values belong to the same category, the confusion matrix marks it an instance of agreement (in this context, CalCAT ensemble values are treated as the source of truth, or actuals). The total frequency of agreement instances for each *R_e_* category are then visualized by the confusion matrix.

We reported the resulting sensitivity, specificity, positive predictive values (PPV), and negative predictive values (NPV) for each *R_e_* category. Sensitivity and specificity reflected the fraction of CalCAT ensemble observations which were correctly classified by *R_ww_* for each *R_e_* category. PPV and NPV reflected the proportion of matching and non-matching classifications for *R_ww_* predictions for each *R_e_* category. Here, for a given *R_e_* category, “true positives” indicate reference observations correctly classified by *R_ww_* as belonging to the category, and “true negatives” indicate reference observations correctly classified by *R_ww_*as not belonging to the category. We also reported the overall accuracy, which summarizes across all *R_e_* categories. The overall accuracy sums the number of *R_e_* predictions correctly classified by *R_ww_* and divides that sum by the total number of observations in the time series. These five metrics do not include the categories *R_e_* < 0.7 and *R_e_*> 1.3 as they had no values during the study period.

We used the R package caret to generate all confusion matrices and corresponding performance metrics.^42^

### Comparison of R_e_ temporalities

We performed cross-correlation between *R_ww_* and *R_cc_* to identify temporal lags in the former that may be useful predictors of the latter. Cross-correlations were performed for a lag value range of -20 to 20 days (negative lag values: *R_ww_* temporally precedes *R_cc_*; positive lag values: *R_cc_* temporally precedes *R_ww_* ; lag value of zero: *R_ww_* is not temporally shifted with respect to *R_cc_*).

## RESULTS

We produced two SARS-CoV-2 *R_ww_* models for five California counties. We then compared the timing, magnitude, and directionality of *R_ww_* to the CalCAT ensemble and sewershed-restricted *R_cc_*. All results for San Francisco, Santa Clara, and Yolo counties are included in the main text; numerical results for Sacramento and Stanislaus counties are included in the main text and graphical results are in the Supplemental Material.

### Comparison of R_e_ trajectories

*R_ww_* tracks closely with *R_cc_* over time and geography (Figures 3, S2, and S4). Quantitatively, we found high correspondence between *R_ww_* and *R_cc_* trajectories for all counties across three analyses: mean absolute error (MAE), Spearman’s rank correlation, and confusion matrix classification.

**Figure 3.**
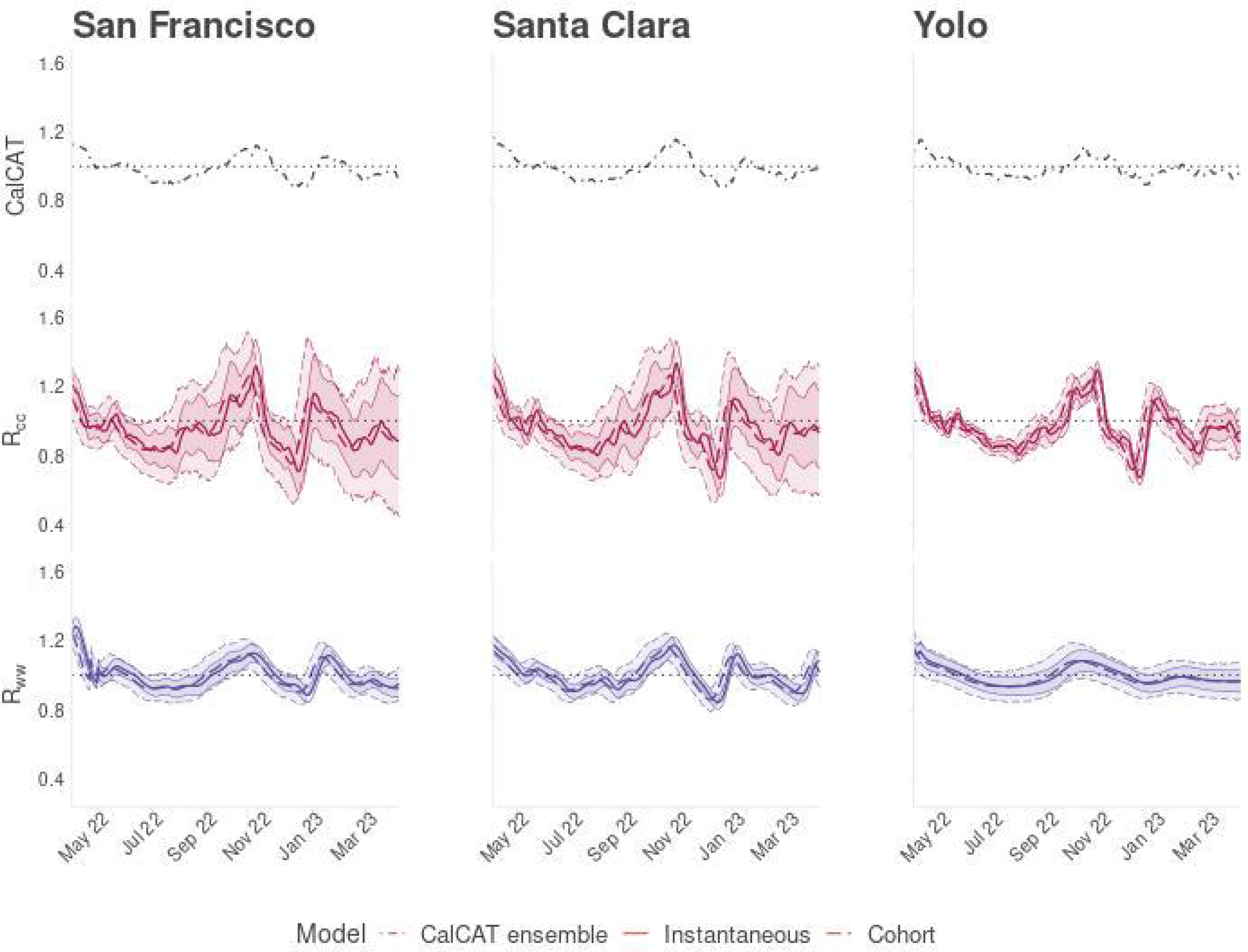
Time series of county-aggregated, sewershed-restricted *R_e_* for Santa Clara, San Francisco, and Yolo. Between May 1, 2022 and April 30, 2023, three county-level *R_e_*time series are compared: (top, black) the CalCAT ensemble – a publicly available ensemble of county-wide *R_cc_* estimates; (middle, pink) county-aggregated, sewershed-restricted *R_cc_*; and (bottom, blue) *R_ww_*. Both *R_ww_* and sewershed-restricted *R_cc_*were calculated using the *R_e_* estimation pipeline piloted in this study. Solid pink or blue lines indicate instantaneous *R_e_*, while dashed pink or blue lines indicate cohort *R_e_*. 95% confidence intervals for each *R_e_* type (instantaneous or cohort) are depicted. Results for three of five studied counties (San Francisco, Santa Clara, and Yolo) are included; results for the remaining counties (Sacramento and Stanislaus) are included in the Supplemental Material. Note: *R_ww_*, sewershed-restricted, wastewater-based effective reproduction number; *R_cc_*, case-based effective reproduction number.

#### Mean Absolute Error Analysis

When compared against *R_cc_* for the entire analysis period, *R_ww_* had a low average MAE (MAE ≤ 0.09, Table 3). For both instantaneous and cohort *R_ww_* models, MAE values relating *R_ww_* to CalCAT ensemble estimates were lower (MAE ≤ 0.07) than MAE values relating *R_ww_* to sewershed-restricted *R_cc_* estimates (MAE ≤ 0.09). MAE values relating *R_ww_* to CalCAT ensemble estimates were lower for instantaneous *R_ww_* (MAE ≤ .05), while MAE values relating *R_ww_* to sewershed-restricted *R_cc_* estimates were lower for cohort *R_ww_*(MAE ≤ 0.06). These results were consistent across all five counties.

**Table 3.** Comparative analyses of county-level *R_ww_* and *R_cc_* trajectories using mean absolute error and Spearman’s rank correlation.

| County | Type of $R_{cc}$ estimate | MAE, Instantaneous $R_{ww}$ | MAE, Cohort $R_{ww}$ | Spearman's $\rho$, Instantaneous $R_{ww}$ | $\rho$ p-value, Instantaneous $R_{ww}$ | Spearman's $\rho$, Cohort $R_{ww}$ | $\rho$ p-value, Cohort $R_{ww}$ |
| --- | --- | --- | --- | --- | --- | --- | --- |
| Sacramento | Sewershed-restricted cases | 0.082 | 0.072 | 0.719 | 3.20E-59 | 0.712 | 8.80E-58 |
|  | CalCAT ensemble | 0.042 | 0.052 | 0.775 | 3.10E-74 | 0.621 | 2.30E-40 |
| San Francisco | Sewershed-restricted cases | 0.077 | 0.071 | 0.726 | 5.30E-61 | 0.754 | 3.10E-68 |
|  | CalCAT ensemble | 0.025 | 0.031 | 0.936 | 3.80E-167 | 0.866 | 2.50E-111 |
| Santa Clara | Sewershed-restricted cases | 0.064 | 0.059 | 0.800 | 1.20E-82 | 0.823 | 5.50E-91 |
|  | CalCAT ensemble | 0.028 | 0.043 | 0.843 | 9.10E-100 | 0.668 | 1.70E-48 |
| Stanislaus | Sewershed-restricted cases | 0.087 | 0.066 | 0.779 | 1.30E-75 | 0.873 | 1.60E-115 |
|  | CalCAT ensemble | 0.052 | 0.065 | 0.775 | 2.20E-74 | 0.662 | 2.50E-47 |
| Yolo | Sewershed-restricted cases | 0.081 | 0.073 | 0.657 | 2.10E-46 | 0.658 | 1.30E-46 |
|  | CalCAT ensemble | 0.016 | 0.019 | 0.891 | 3.20E-126 | 0.884 | 1.10E-121 |

Note: By calculating mean absolute errors and Spearman's rank correlations, we compared the magnitude and direction of instantaneous and cohort $R_{ww}$ models to two types of $R_{cc}$ estimates: (1) a publicly available ensemble of county-wide $R_{cc}$ estimates ("CalCAT ensemble"), and (2) a county-aggregated, sewershed-restricted $R_{cc}$ . Reported p-values correspond to Spearman's rank correlation, $\rho$ ( $R_{ww}$ compared against $R_{cc}$ ). Abbreviations: $R_{ww}$ , sewershed-restricted, wastewater-based effective reproduction number; $R_{cc}$ , case-based effective reproduction number; MAE, mean absolute error; $\rho$ (rho), Spearman's rank correlation coefficient.

#### Spearman’s Rank Correlation Analysis

Over the entire analysis period, *R_ww_* estimates were strongly, positively, and significantly correlated with *R_cc_* estimates for all five counties (Spearman ρ ≥ 0.62, p < 0.001) (Table 3). For instantaneous *R_ww_*, strength of correlation between *R_ww_* and CalCAT ensemble estimates was higher (Spearman ρ ≥ 0.78, p < 0.001) than the strength of correlation between *R_ww_* and sewershed-restricted *R_cc_* estimates (Spearman ρ ≥ 0.66, p < 0.001) in four of five counties. For cohort *R_ww_*, strength of correlation between *R_ww_* and the CalCAT ensemble (Spearman ρ ≥ 0.62, p < 0.001) did not show a clear pattern in relation to the strength of correlation between *R_ww_* and sewershed-restricted *R_cc_* estimates (Spearman ρ ≥ 0.66, p < 0.001). Strength of correlation between *R_ww_* and the CalCAT ensemble estimates was higher across all counties for the instantaneous *R_ww_*model (Spearman ρ ≥ 0.78, p < 0.001). The strength of correlation between *R_ww_* and sewershed-restricted *R_cc_* estimates was generally higher for the cohort *R_ww_* approach (Spearman ρ ≥ 0.62, p < 0.001), with the exceptions of Sacramento and Yolo counties (for which instantaneous *R_ww_* demonstrated slightly higher strength of correlation).

#### Confusion Matrix Analysis

We performed confusion matrix classification analysis, stratified by county and *R_ww_*modeling approach, to further characterize the directional concordance between *R_ww_* and the CalCAT ensemble (Figures 4 and S3).

**Figure 4.**
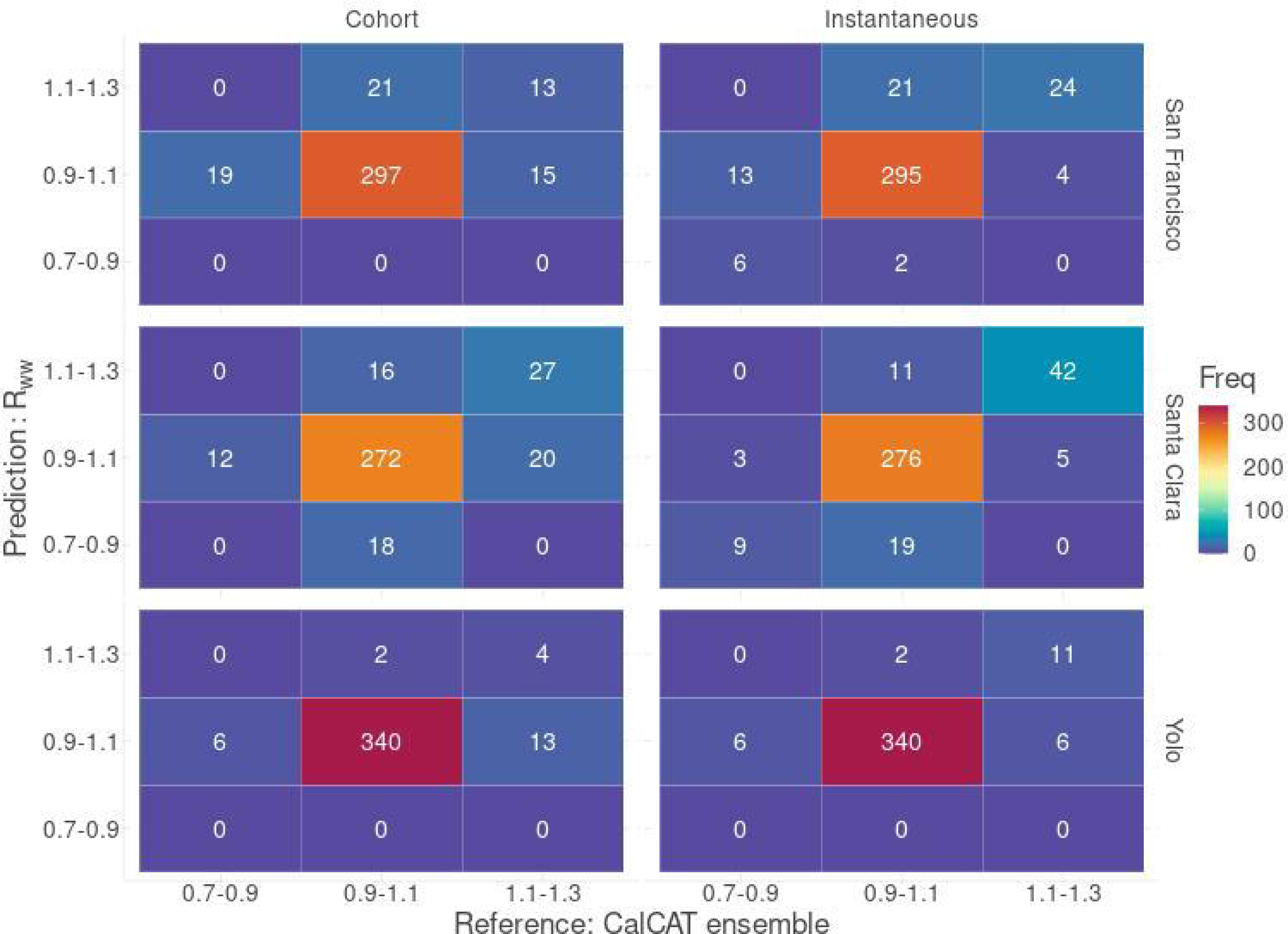
Frequency of agreement between *R_ww_* and the CalCAT ensemble for Santa Clara, San Francisco and Yolo. Based on magnitude, *R_e_* values were classified into transmission strength categories (<0.7-0.9, 0.9-1.1, 1.1-1.3, >1.3, which represent a strong decrease, decrease, stability, increase, and strong increase in *R_e_*, respectively). Frequency of agreement between *R_ww_* and the CalCAT ensemble (i.e., instances when predicted *R_ww_* values and reference CalCAT ensemble values belong to the same *R_e_* category) are visualized by the confusion matrix. The right column illustrates results for cohort *R_ww_*, and the left for instantaneous *R_ww_*. Each row represents a single county. The counter diagonals (top right to bottom left) of each matrix represents true positives. Off-diagonal values indicate instances of disagreement between *R_ww_* model predictions and the CalCAT ensemble. Two *R_e_* categories (*R_e_* < 0.7, *R_e_* >1.3) with no *R_e_*values during the study period were excluded. Results for three of five studied counties (San Francisco, Santa Clara, and Yolo) are included; results for the remaining counties (Sacramento and Stanislaus) are included in the Supplemental Material. Note: *R_ww_*, sewershed-restricted, wastewater-based effective reproduction number; Instantaneous, instantaneous *R_ww_*; Cohort, cohort *R_ww_*.

The overall classification accuracy of both *R_ww_* models was consistently high (≥ 0.79) across all counties; the sole exception to this pattern was Stanislaus, which had a slightly lower overall accuracy for its *R_ww_* models (≥ 0.64) (Tables 4 and 5). Overall accuracy for the cohort *R_ww_* model was lower as compared to instantaneous *R_ww_*for all counties. For all counties and *R_ww_* approaches, the total frequency of agreement instances was greatest for the 0.9 to 1.1 *R_e_* (stable) range (Figures 4 and S3).

**Table 4.** Summary metrics from multi-class confusion matrix analysis evaluating agreement between instantaneous *R_ww_* and the CalCAT ensemble.

| County | $R_e$<br>Classification | Overall<br>Accuracy | Sensitivity | Specificity | PPV | NPV |
| --- | --- | --- | --- | --- | --- | --- |
| Sacramento | 0.7-0.9 | 0.87 | 0.73 | 0.89 | 0.17 | 0.99 |
|  | 0.9-1.1 | <sup>a</sup> | 0.87 | 0.84 | 0.97 | 0.51 |
|  | 1.1-1.3 |  | 0.87 | 0.99 | 0.94 | 0.98 |
| San Francisco | 0.7-0.9 | 0.89 | 0.32 | 0.99 | 0.75 | 0.96 |
|  | 0.9-1.1 |  | 0.93 | 0.64 | 0.95 | 0.57 |
|  | 1.1-1.3 |  | 0.86 | 0.94 | 0.53 | 0.99 |
| Santa Clara | 0.7-0.9 | 0.90 | 0.75 | 0.95 | 0.32 | 0.99 |
|  | 0.9-1.1 |  | 0.90 | 0.86 | 0.97 | 0.63 |
|  | 1.1-1.3 |  | 0.89 | 0.97 | 0.79 | 0.98 |
| Stanislaus | 0.7-0.9 | 0.76 | 0.95 | 0.94 | 0.73 | 0.99 |
|  | 0.9-1.1 |  | 0.78 | 0.79 | 0.89 | 0.63 |
|  | 1.1-1.3 |  | 0.52 | 0.88 | 0.46 | 0.9 |
| Yolo | 0.7-0.9 | 0.96 | 0 | 1 | <sup>b</sup>NA | 0.98 |
|  | 0.9-1.1 |  | 0.99 | 0.48 | 0.97 | 0.85 |
|  | 1.1-1.3 |  | 0.65 | 0.99 | 0.85 | 0.98 |

Note: We produced multi-class confusion matrices relating instantaneous $R_{ww}$ to the CalCAT ensemble (real-time, ongoing $R_{cc}$ estimates with relevance for statewide public health). $R_e$ values were classified into transmission strength categories based on magnitude (<0.7-0.9, 0.9-1.1, 1.1-1.3, >1.3, which represent a strong decrease, decrease, stability, increase, and strong increase in transmission, respectively). The resulting sensitivity, specificity, positive predictive and negative predictive values for each $R_e$ category are reported. The overall accuracy for each county-level $R_{ww}$ is also reported. We did not report two $R_e$ categories (< 0.7, >1.3), which had no observations during the study period. <sup>a</sup>All gray boxes indicate values identical to the closest, previous non-zero entry within the same column. <sup>b</sup>NA entries correspond to $R_e$ categories with zero $R_{ww}$ predictions during the study period. $R_e$ , effective reproduction number; PPV, Positive Predictive Value; NPV, Negative Predictive Value.

**Table 5.** Summary metrics from multi-class confusion matrix analysis evaluating agreement between cohort *R_ww_* and the CalCAT ensemble.

| County | $R_e$<br>Classification | Overall<br>Accuracy | Sensitivity | Specificity | PPV | NPV |
| --- | --- | --- | --- | --- | --- | --- |
| Sacramento | 0.7-0.9 | 0.79 | 0 | 0.87 | 0 | 0.97 |
|  | 0.9-1.1 | <sup>a</sup> | 0.84 | 0.46 | 0.91 | 0.31 |
|  | 1.1-1.3 |  | 0.59 | 0.98 | 0.82 | 0.95 |
| San Francisco | 0.7-0.9 | 0.85 | 0 | 1 | <sup>b</sup>NA | 0.95 |
|  | 0.9-1.1 |  | 0.93 | 0.28 | 0.9 | 0.38 |
|  | 1.1-1.3 |  | 0.46 | 0.94 | 0.38 | 0.95 |
| Santa Clara | 0.7-0.9 | 0.82 | 0 | 0.95 | 0 | 0.97 |
|  | 0.9-1.1 |  | 0.89 | 0.46 | 0.89 | 0.44 |
|  | 1.1-1.3 |  | 0.57 | 0.95 | 0.63 | 0.94 |
| Stanislaus | 0.7-0.9 | 0.64 | 0.54 | 0.89 | 0.48 | 0.91 |
|  | 0.9-1.1 |  | 0.70 | 0.56 | 0.77 | 0.47 |
|  | 1.1-1.3 |  | 0.5 | 0.86 | 0.41 | 0.9 |
| Yolo | 0.7-0.9 | 0.94 | 0 | 1 | NA | 0.98 |
|  | 0.9-1.1 |  | 0.99 | 0.17 | 0.95 | 0.67 |
|  | 1.1-1.3 |  | 0.24 | 0.99 | 0.67 | 0.96 |

Note: We produced multi-class confusion matrices relating instantaneous $R_{ww}$ to the CalCAT ensemble (real-time, ongoing $R_{cc}$ estimates with relevance for statewide public health). $R_e$ values were classified into transmission strength categories based on magnitude (<0.7-0.9, 0.9-1.1, 1.1-1.3, >1.3, which represent a strong decrease, decrease, stability, increase, and strong increase in $R_e$ , respectively). The resulting sensitivity, specificity, positive predictive and negative predictive values for each $R_e$ category are reported. The overall accuracy for each county-level $R_{ww}$ is also reported. We did not report two $R_e$ categories (< 0.7, >1.3), which had no observations during the study period. <sup>a</sup>All gray boxes indicate values identical to the closest, previous non-zero entry within the same column. <sup>b</sup>NA entries correspond to $R_e$ categories with zero $R_{ww}$ values during the study period. $R_e$ , effective reproduction number; PPV, Positive Predictive Value; NPV, Negative Predictive Value.

Across all counties, both *R_ww_* models generally demonstrated the greatest sensitivity and PPV for *R_e_* estimates with magnitudes between 0.9 and 1.1 (sensitivity = 0.78-0.99 and 0.70-0.99; PPV = 0.89-0.97 and 0.77-0.95 for instantaneous and cohort, respectively). For all *R_e_*classifications, sensitivity and PPV varied by geography and *R_ww_*model type. Interestingly, for *R_e_* estimates between 0.7 and 0.9, the PPV of cohort *R_ww_* was incalculable (San Francisco and Yolo) or 0 (Sacramento and Santa Clara) for all counties except Stanislaus (PPV = 0.48). An incalculable PPV indicates there were no *R_ww_* predictions within this range; a PPV of 0 indicates *R_ww_* never estimated *R_e_* between 0.7 and 0.9 when the CalCAT ensemble did. However, it is important to caveat that there were few (< 20) eligible observations in this category for all counties except Stanislaus.

For *R_e_* estimates between 0.7 and 0.9 or 1.1 and 1.3, all misclassified *R_e_* (i.e., *R_e_* estimates categorized differently by the CalCAT ensemble versus *R_ww_*) were estimated to be in the 0.9 to 1.1 range by both *R_ww_* models. Consistent with this finding, both *R_ww_* models demonstrated the lowest specificity and NPV for the 0.9 to 1.1 category across all counties (specificity = 0.48-0.86 and 0.17-0.56; NPV = 0.51-0.85 and 0.31-0.67 for instantaneous and cohort, respectively). For *R_e_* estimates between 0.7 and 0.9 or 1.1 and 1.3, the specificity and NPV remained greater than 0.86 and 0.90, respectively, across all counties for both *R_ww_* models. We observed inter- and intra-county- and model-dependent variability in relative magnitudes of the four reported metrics (sensitivity, specificity, PPV, NPV) across *R_e_* classifications.

### Comparison of R_e_ temporalities

We calculated cross-correlation with a maximum lag of 20 days between *R_ww_* and *R_cc_* to numerically evaluate temporal alignment. Across all counties and *R_ww_*-*R_cc_* pairings, cross-correlation coefficients ranged from 0.65 to 0.93 (CalCAT ensemble: 0.72-0.93 and 0.69-0.89; sewershed-restricted *R_cc_*: 0.65-0.87 and 0.67-0.89 for instantaneous and cohort *R_ww_*, respectively). We report the range of time lags for cross-correlation coefficients within 0.05 of the maximum observed correlation value (Table 6, Figures S5 and S6). The reported range of time lags were wide and variable across counties, *R_cc_* sources (CalCAT ensemble versus sewershed-restricted), and *R_ww_* models (instantaneous versus cohort). Importantly, though, nearly all ranges included a lead time of 0 days, when *R_ww_* was not temporally shifted with respect to *R_cc_* (the two exceptions to this were: (1) instantaneous *R_ww_* with respect to sewershed-restricted *R_cc_*for Stanislaus county, and (2) cohort *R_ww_* with respect to the CalCAT ensemble for Sacramento county); this is consistent with our previous observations of high concordance between non-time-shifted *R_ww_* and *R_cc_* (Figures 3 and S2, Table 3).

**Table 6.** Comparison of R_e_ temporality via cross correlations.

| County | $R_{cc}$ | Lag (Days),<br>Instantaneous $R_{ww}$ | Lag (Days),<br>Cohort $R_{ww}$ | CC,<br>Instantaneous $R_{ww}$ | CC,<br>Cohort $R_{ww}$ |
| --- | --- | --- | --- | --- | --- |
| Sacramento | Sewershed-restricted cases | [-3, 4] | [-4, 3] | [0.65, 0.7] | [0.67, 0.72] |
|  | CalCAT ensemble | [-8, 2] | [-15, -4] | [0.72, 0.77] | [0.69, 0.73] |
| San Francisco | Sewershed-restricted cases | [0, 8] | [0, 10] | [0.73, 0.77] | [0.76, 0.80] |
|  | CalCAT ensemble | [-4, 2] | [-11, 0] | [0.87, 0.92] | [0.84, 0.89] |
| Santa Clara | Sewershed-restricted cases | [-2, 3] | [-3, 4] | [0.84, 0.87] | [0.84, 0.89] |
|  | CalCAT ensemble | [-8, 1] | [-16, -6] | [0.84, 0.88] | [0.8, 0.84] |
| Stanislaus | Sewershed-restricted cases | [4, 10] | [-1, 7] | [0.72, 0.76] | [0.81, 0.85] |
|  | CalCAT ensemble | [-6, 2] | [-15, -6] | [0.73, 0.76] | [0.78, 0.83] |
| Yolo | Sewershed-restricted cases | [-2, 9] | [-2, 10] | [0.68, 0.73] | [0.68, 0.73] |
|  | CalCAT ensemble | [-5, 3] | [-9, 1] | [0.88, 0.93] | [0.84, 0.89] |

Note: Using cross-correlation analysis with a maximum lag of 20 days, we investigated temporal alignment of $R_{ww}$ (cohort or instantaneous) and $R_{cc}$ . $R_{cc}$ included either the CalCAT ensemble – a publicly available ensemble of county-wide $R_{cc}$ estimates – or county-aggregated, sewershed-restricted $R_{cc}$ estimates. We report the range of time lags for cross-correlation coefficients within 0.05 of the maximum observed correlation value. We also include the range of cross-correlation coefficients within 0.05 of the maximum observed correlation value. Negative lag values indicate $R_{ww}$ temporally precedes $R_{cc}$ ; positive lag values indicate $R_{cc}$ temporally precedes $R_{ww}$ ; lag values of zero indicate no temporal shift of $R_{ww}$ with respect to $R_{cc}$ . $R_{ww}$ , sewershed-restricted, wastewater-based effective reproduction number; $R_{cc}$ , case-based effective reproduction number; CC, cross-correlation coefficient

Within each county, the size and bounds (i.e., minimum and maximum) of the reported range of time lags with respect to sewershed-restricted *R_cc_* was broadly similar between both *R_ww_* models; this was not true for the range of time lags with respect to the CalCAT ensemble, which differs between *R_ww_* models (boundaries of ranges with respect to the CalCAT ensemble: -3 to +10 days and -4 to +10 days of lag for instantaneous and cohort, respectively; boundaries of ranges with respect to the CalCAT ensemble: -8 to +3 days and -16 to +1 days of lag for instantaneous and cohort, respectively).

## DISCUSSION

We estimated instantaneous and cohort wastewater-based SARS-CoV-2 effective reproduction numbers for five California counties from May 1, 2022 to April 30, 2023. Across counties with varying population characteristics (e.g., demographics, sizes, clinical testing rates) and wastewater surveillance (e.g., sampling frequency, coverage), both instantaneous and cohort *R_ww_* models demonstrated high correspondence with traditional *R_cc_* models based on sewershed-restricted and county-wide case data. Correspondence was indicated by low average MAE, significant positive Spearman correlation, and high classification accuracy of *R_ww_* with respect to *R_cc_* for the entire study period (Table 3, Figure 4). These findings align with existing studies on *R_ww_* estimation, which consistently reveal high concordance between *R_ww_* and *R_cc_* estimates across geographies and time.^17,20,21^ *R_ww_* concordance with the county-level CalCAT ensemble was generally greater than with sewershed-restricted *R_cc_* (Table 3).

Our work did not yield clear conclusions on the relative timing of *R_ww_* and *R_cc_*. Cross correlative analyses suggested strong associations ranging from a lead time of -16 days to a lag time of +10 days, depending on county, *R_cc_* source (CalCAT ensemble versus sewershed-restricted), and *R_ww_* model type (Table 6). These findings align mixed results in previous studies assessing the temporal alignment between wastewater and clinical surveillance data^43–46;^ several factors, such as clinical data completeness, SARS-CoV-2 variant predominance, sewershed location, and population immunity, may influence the strength of wastewater as a leading indicator of disease incidence.

Similar SARS-CoV-2 *R_ww_* estimation pipelines are described in Huisman et al.,^17^ Amman et al.,^19^ and the publicly available “COVID-19 R estimation for California” dashboard by Worden et al. (https://ca-covid-r.info/). Both published studies produce sewershed-level instantaneous *R_ww_*, while the dashboard produces county-level cohort *R_ww_*. All methods share a general pipeline schema: wastewater time series data is first processed and subsequently fed into an *R_e_* model. However, the methods differ in their strategies for data transformation, deconvolution (including selection of infection to shedding delay distributions), and county aggregation. For data transformation, our method is most similar to Amman et al.,^19^ as both implement a spline approach to smooth wastewater data. For deconvolution, our method is most similar to Huisman et al.,^17^ as both implement a variant of the Richardson-Lucy expectation-maximization algorithm. For county aggregation, our method differs from that of the “COVID-19 R estimation for California” dashboard (https://ca-covid-r.info/): we perform population weighting of sewershed-level *R_ww_*, while the dashboard uses non-population-weighted, county-aggregated input wastewater data to estimate *R_ww_*. We chose population-weighting to ensure sewersheds corresponding to larger or smaller populations have proportional influence on and representation in the county-level *R_ww_* estimate. We opted for aggregating sewershed-restricted *R_ww_* instead of sewershed-restricted raw RNA concentrations because the latter are not necessarily comparable between sewersheds due to environmental conditions and wastewater attributes.

There are several limitations to our study. First and most importantly, to validate *R_ww_* we treated *R_cc_* as the gold standard *R_e_*metric. However, both clinical and wastewater data streams have their respective strengths and limitations. Clinical data streams are influenced by heterogeneous, time- and space-varying testing-related factors, ultimately capturing only a subset of infections. Wastewater data streams, while independent of some of the biases impacting clinical surveillance, require accurate characterization of the fecal shedding load distribution in order to be interpreted; the temporal dynamics of fecal shedding and its relationship to variant, immunity status, or disease severity, continue to be an ongoing area of investigation.^47–49^ Given this uncertainty surrounding fecal shedding, pooled viral concentrations in wastewater are difficult to directly translate into unique infection or transmission events. Consequently, *R_ww_* may not truly represent the average number of secondary cases caused by a newly infectious individual. Instead, it may be more indicative of other disease characteristics such as infectiousness (since highest shedding tends to occur when someone is most infectious). This is in contrast to traditional *R_cc_*estimates, which use discrete units of disease burden (e.g., case counts) in conjunction with generation time or serial interval distributions to directly reconstruct and quantify transmission events. Ultimately, these limitations suggest that both clinical and wastewater data streams are imperfect proxies for COVID-19 disease dynamics; together though, both sources could potentially offer a more comprehensive picture of transmission dynamics and infection risk. While the present study frames *R_cc_* as a gold-standard metric, *R_cc_* and *R_ww_* should instead be considered as two complementary *R_e_* metrics informed by synergistic data sources.

Second, county-level *R_ww_* is based on sewershed-restricted wastewater data. Such data only reflects the shedding patterns and disease conditions of wastewater-surveilled communities and may not be generalizable to the entire county population. This has important implications for the utility of *R_ww_* in the context of public health policies, such as health mandates, which are often applied at the county level. Any county-level policies informed by *R_ww_* would be tied to a transmission indicator that can only account for the sewershed-surveilled subset of the infected population. This is especially relevant for the state of California, where most counties (especially those in rural areas) have <50% of the county population surveilled by wastewater. Given that our study focuses on counties with >50% coverage, our findings on the agreement between county-aggregated, sewershed-level *R_ww_* and county-level CalCAT ensemble *R_cc_* may not translate to communities with lower programmatic wastewater surveillance. In addition, the wastewater-surveilled subset may demonstrate unique behaviors that are not representative of the entire county, potentially biasing *R_ww_*.

Third, wastewater data is susceptible to location- or time-specific environmental conditions. Wastewater samples are relatively small volumes of wastewater collected from millions of gallons of organic and inorganic materials flowing through a treatment plant; as such, factors like rainfall, industrial input, temperature, and sewage travel time can impact measured SARS-CoV-2 RNA concentrations. This may weaken *R_ww_* –*R_cc_* concordance in specific counties.

There are also key limitations within our *R_ww_* estimation process. For all five counties in our study, we assume the experimentally inferred, optimized infection to shedding distribution for San Jose described by Huisman et al.^17^ This distribution maximized *R_ww_*-*R_cc_* agreement for a single treatment plant studied by Huisman et al.,^17^ and is therefore not reflective of the highly variable fecal shedding profile amongst individuals and populations. This upstream assumption in our pipeline impacts the number of inferred infections per day from wastewater, ultimately influencing *R_ww_* estimates. Similarly, specific decisions were made in the estimation process related to smoothing (e.g., methodology), trimming (e.g., width or window), and deconvolution (e.g., choice of incubation period, generation time) which may influence downstream results.

Lastly, it is important to consider practical limitations relevant for operationalization of our methods. Firstly, our study is a retrospective analysis using readily available historical surveillance data that has been updated post hoc as more accurate information was reported. This contrasts with real-time or emergency conditions, when both *R_cc_* and *R_ww_* can only rely on data available at a given time. As such, real-time operationalization of the *R_ww_* estimation procedure outlined in this study may require additional data pre-processing, nowcasting, and forecasting steps in the absence of current data. Secondly, counties were selected for this pilot analysis to prioritize similar laboratorial methodologies, long-term sampling histories, high population coverage by wastewater surveillance, and representation of diverse (rural and urban) demographics. Such historically comprehensive, consistently measured, and frequently sampled wastewater data is not available for all sewersheds, counties, or regions. Therefore, the performance of the *R_ww_* estimation process presented here may vary considerably when applied to other jurisdictions in California and may not be generalizable. In the future, investigating *R_ww_* estimation in a larger suite of counties and regions will be essential to comprehensively understanding the utility of our method in a wide range of scenarios. Another future extension includes identifying the minimum wastewater county coverage and sampling frequency required for high *R_ww_*-*R_cc_* concordance, which would increase county representation (building upon initial work done by Huisman et al. 2022 for San Jose). Finally, methodological differences can impact measured wastewater concentrations, which in turn can complicates comparability between sites. While we selected sewersheds using similar laboratory methods for this study, some differences still remained in how wastewater solids were collected, processed, or quantified. We do not have conclusive evidence on the impact of even minor differences in laboratory methodologies on downstream *R_ww_* estimation. Moreover, best practices for combining wastewater data across multiple methods and laboratories (either over time for a single site, or for distinct sites within a single county) remain an area of ongoing and future research.

We estimated county-level *R_ww_*, which we demonstrated tracks closely with robust traditional case-based *R_cc_*estimates. Our results support the future use of *R_ww_* as an additional epidemiological tool with public health relevance in the context of disease dynamics monitoring. Moreover, our study provides a generalizable, robust, and operationalizable framework for estimating county-level *R_ww_*. On the basis of this research, we produced publicly available SARS-CoV-2 *R_ww_*nowcasts for the CalCAT dashboard (https://calcat.covid19.ca.gov/cacovidmodels/), advancing COVID-19 transmission monitoring for the state of California. This study and the described approach can inform other jurisdictions’ construction and implementation of sewershed- and county-level *R_ww_* estimation pipelines. In the future, the described estimation procedure could be applied to other pathogens with available wastewater and case surveillance data (e.g., respiratory syncytial virus [RSV], influenza).

## Data Availability

All wastewater data used in this study is publicly accessible through the CA Open Data Portal at https://data.ca.gov/dataset/covid-19-wastewater-surveillance-data-california. Models contributing to the CalCAT ensemble also use data publicly accessible through the CA Open Data Portal, such as downloadable case counts, hospitalizations, deaths and test positivity (https://data.chhs.ca.gov/organization/california-department-of-public-health). Sewershed-restricted COVID-19 case data reported to CDPH is not publicly available; readers should contact the corresponding author with data requests.

https://data.chhs.ca.gov/organization/california-department-of-public-health

## ACKNOWLEDGEMENTS

The wastewater sampling conducted through Method 1 was supported by gifts from the CDC Foundation and the Sergey Brin Family Foundation to ABB. Additional research support for Method 2 were provided through a philanthropic gift to the University of California, Davis. CDPH wastewater surveillance efforts are supported in part by the Epidemiology and Laboratory Capacity for Infectious Diseases Cooperative Agreement (no. 6NU50CK000539-04-02) from CDC.

The authors sincerely thank the operators at Sacramento Regional Wastewater Treatment Plan, Esparto Wastewater Treatment Facility, Woodland Water Pollution Control Facility, Winters Water Treatment Facility, City of Davis Wastewater Treatment Plant, Modesto’s Sutter Primary Treatment Facility, Turlock Regional Water Quality Control Facility, Silicon Valley Clean Water, San Jose Santa Clara Regional Wastewater Facility, City of Sunnyvale Water Pollution Control Plant, Palo Alto Regional Water Quality Control Plant, South County Regional Wastewater Authority, Oceanside Water Pollution Control Plant, and Southeast Water Pollution Control Plant for providing wastewater samples.

The authors further thank the modeling and wastewater surveillance sections at CDPH for input and guidance on the data sources and procedure for *R_ww_*estimation outlined in this study.

## PUBLICATION POLICY DISCLAIMER

The findings and conclusions in this article are those of the author(s) and do not necessarily represent the views or opinions of the California Department of Public Health or the California Health and Human Services Agency.

## Notes

### Author Declarations

The Committee for the Protection of Human Subjects (CPHS) of the California Health and Human Services waived ethical approval for this work. CPHS determined that our proposed project is exempt under their criteria.

## REFERENCES

1. New Framework and Software to Estimate Time-Varying Reproduction Numbers During Epidemics | American Journal of Epidemiology | Oxford Academic. Accessed November 21, 2023. https://academic.oup.com/aje/article/178/9/1505/89262?login=false

2. Joint Research Centre (European Commission), Annunziato A, Asikainen T. Effective Reproduction Number Estimation from Data Series. Publications Office of the European Union; 2020. Accessed November 21, 2023. https://data.europa.eu/doi/10.2760/036156

3. Linka K, Peirlinck M, Kuhl E. The reproduction number of COVID-19 and its correlation with public health interventions. Comput Mech. 2020;66(4):1035–1050. doi:10.1007/s00466-020-01880-8

4. Fauci AS, Lane HC, Redfield RR. Covid-19 — Navigating the Uncharted. N Engl J Med. 2020;382(13):1268–1269. doi:10.1056/NEJMe2002387

5. Dainton C, Hay A. Quantifying the relationship between lockdowns, mobility, and effective reproduction number (Rt) during the COVID-19 pandemic in the Greater Toronto Area. BMC Public Health. 2021;21(1):1658. doi:10.1186/s12889-021-11684-x

6. Inglesby TV. Public Health Measures and the Reproduction Number of SARS-CoV-2. JAMA. 2020;323(21):2186–2187. doi:10.1001/jama.2020.7878

7. Pitzer VE, Chitwood M, Havumaki J, et al. The Impact of Changes in Diagnostic Testing Practices on Estimates of COVID-19 Transmission in the United States. Am J Epidemiol. Published online April 8, 2021:kwab089. doi:10.1093/aje/kwab089

8. Nash RK, Nouvellet P, Cori A. Real-time estimation of the epidemic reproduction number: Scoping review of the applications and challenges. PLOS Digit Health. 2022;1(6):e0000052. doi:10.1371/journal.pdig.0000052

9. Li X, Zhang S, Sherchan S, et al. Correlation between SARS-CoV-2 RNA concentration in wastewater and COVID-19 cases in community: A systematic review and meta-analysis. J Hazard Mater. 2023;441:129848. doi:10.1016/j.jhazmat.2022.129848

10. Rabe A, Ravuri S, Burnor E, et al. Correlation between wastewater and COVID-19 case incidence rates in major California sewersheds across three variant periods. J Water Health. 2023;21(9):1303–1317. doi:10.2166/wh.2023.173

11. Bivins A, North D, Ahmad A, et al. Wastewater-Based Epidemiology: Global Collaborative to Maximize Contributions in the Fight Against COVID-19. Environ Sci Technol. 2020;54(13):7754–7757. doi:10.1021/acs.est.0c02388

12. Peccia J, Zulli A, Brackney DE, et al. Measurement of SARS-CoV-2 RNA in wastewater tracks community infection dynamics. Nat Biotechnol. 2020;38(10):1164–1167. doi:10.1038/s41587-020-0684-z

13. Wolfe MK, Topol A, Knudson A, et al. High-Frequency, High-Throughput Quantification of SARS-CoV-2 RNA in Wastewater Settled Solids at Eight Publicly Owned Treatment Works in Northern California Shows Strong Association with COVID-19 Incidence. mSystems. 2021;6(5):10.1128/msystems.00829-21. doi:10.1128/msystems.00829-21

14. Kirby AE, Walters MS, Jennings WC, et al. Using Wastewater Surveillance Data to Support the COVID-19 Response — United States, 2020–2021. Morb Mortal Wkly Rep. 2021;70(36):1242–1244. doi:10.15585/mmwr.mm7036a2

15. Olesen SW, Imakaev M, Duvallet C. Making waves: Defining the lead time of wastewater-based epidemiology for COVID-19. Water Res. 2021;202:117433. doi:10.1016/j.watres.2021.117433

16. Zhu Y, Oishi W, Maruo C, et al. Early warning of COVID-19 via wastewater-based epidemiology: potential and bottlenecks. Sci Total Environ. 2021;767:145124. doi:10.1016/j.scitotenv.2021.145124

17. Huisman JS, Scire J, Caduff L, et al. Wastewater-Based Estimation of the Effective Reproductive Number of SARS-CoV-2. Environ Health Perspect. 2022;130(5):057011. doi:10.1289/EHP10050

18. Nourbakhsh S, Fazil A, Li M, et al. A wastewater-based epidemic model for SARS-CoV-2 with application to three Canadian cities. Epidemics. 2022;39:100560. doi:10.1016/j.epidem.2022.100560

19. Amman F, Markt R, Endler L, et al. Viral variant-resolved wastewater surveillance of SARS-CoV-2 at national scale. Nat Biotechnol. 2022;40(12):1814–1822. doi:10.1038/s41587-022-01387-y

20. Nadeau S, Devaux AJ, Bagutti C, et al. Influenza transmission dynamics quantified from wastewater. Published online January 25, 2023:2023.01.23.23284894. doi:10.1101/2023.01.23.23284894

21. Jiang G, Wu J, Weidhaas J, et al. Artificial neural network-based estimation of COVID-19 case numbers and effective reproduction rate using wastewater-based epidemiology. Water Res. 2022;218:118451. doi:10.1016/j.watres.2022.118451

22. Wallinga J, Teunis P. Different epidemic curves for severe acute respiratory syndrome reveal similar impacts of control measures. Am J Epidemiol. 2004;160(6):509–516. doi:10.1093/aje/kwh255

23. Borchardt MA, Boehm AB, Salit M, Spencer SK, Wigginton KR, Noble RT. The Environmental Microbiology Minimum Information (EMMI) Guidelines: qPCR and dPCR Quality and Reporting for Environmental Microbiology. Environ Sci Technol. 2021;55(15):10210–10223. doi:10.1021/acs.est.1c01767

24. Kim S, C. Kennedy L, K. Wolfe M, et al. SARS-CoV-2 RNA is enriched by orders of magnitude in primary settled solids relative to liquid wastewater at publicly owned treatment works. Environ Sci Water Res Technol. 2022;8(4):757–770. doi:10.1039/D1EW00826A

25. Wolfe MK, Archana A, Catoe D, et al. Scaling of SARS-CoV-2 RNA in Settled Solids from Multiple Wastewater Treatment Plants to Compare Incidence Rates of Laboratory-Confirmed COVID-19 in Their Sewersheds. Environ Sci Technol Lett. 2021;8(5):398–404. doi:10.1021/acs.estlett.1c00184

26. Boehm AB, Wolfe MK, Wigginton KR, et al. Human viral nucleic acids concentrations in wastewater solids from Central and Coastal California USA. Sci Data. 2023;10(1):396. doi:10.1038/s41597-023-02297-7

27. Kadonsky KF, Naughton CC, Susa M, et al. Expansion of wastewater-based disease surveillance to improve health equity in California’s Central Valley: sequential shifts in case-to-wastewater and hospitalization-to-wastewater ratios. Front Public Health. 2023;11. Accessed November 21, 2023. https://www.frontiersin.org/articles/10.3389/fpubh.2023.1141097

28. M. Pecson B, Darby E, N. Haas C, et al. Reproducibility and sensitivity of 36 methods to quantify the SARS-CoV-2 genetic signal in raw wastewater: findings from an interlaboratory methods evaluation in the U.S. Environ Sci Water Res Technol. 2021;7(3):504–520. doi:10.1039/D0EW00946F

29. Medina CY, Kadonsky KF, Roman FA, et al. The need of an environmental justice approach for wastewater based epidemiology for rural and disadvantaged communities: A review in California. Curr Opin Environ Sci Health. 2022;27:100348. doi:10.1016/j.coesh.2022.100348

30. Yu AT, Hughes B, Wolfe MK, et al. Estimating Relative Abundance of 2 SARS-CoV-2 Variants through Wastewater Surveillance at 2 Large Metropolitan Sites, United States. Emerg Infect Dis. 2022;28(5):940–947. doi:10.3201/eid2805.212488

31. White LA, McCorvie R, Crow D, Jain S, León TM. Assessing the accuracy of California county level COVID-19 hospitalization forecasts to inform public policy decision making. BMC Public Health. 2023;23(1):782. doi:10.1186/s12889-023-15649-0

32. Evaluation of individual and ensemble probabilistic forecasts of COVID-19 mortality in the United States | PNAS. Accessed November 21, 2023. https://www.pnas.org/doi/abs/10.1073/pnas.2113561119

33. Helwig N. npreg: Nonparametric Regression via Smoothing Splines. Published online 2022. https://CRAN.R-project.org/package=npreg

34. Gostic KM, McGough L, Baskerville EB, et al. Practical considerations for measuring the effective reproductive number, Rt. PLOS Comput Biol. 2020;16(12):e1008409. doi:10.1371/journal.pcbi.1008409

35. Del Águila-Mejía J, Wallmann R, Calvo-Montes J, Rodríguez-Lozano J, Valle-Madrazo T, Aginagalde- Llorente A. Secondary Attack Rate, Transmission and Incubation Periods, and Serial Interval of SARS-CoV-2 Omicron Variant, Spain. Emerg Infect Dis. 2022;28(6):1224–1228. doi:10.3201/eid2806.220158

36. Scire J, Huisman JS, Grosu A, et al. estimateR: an R package to estimate and monitor the effective reproductive number. BMC Bioinformatics. 2023;24(1):310. doi:10.1186/s12859-023-05428-4

37. Pierre-Yves Boelle, Obadia T. R0: Estimation of R0 and Real-Time Reproduction Number from Epidemics. Published online 2023. https://CRAN.R-project.org/package=R0

38. Cauchemez S, Boëlle PY, Thomas G, Valleron AJ. Estimating in Real Time the Efficacy of Measures to Control Emerging Communicable Diseases. Am J Epidemiol. 2006;164(6):591–597. doi:10.1093/aje/kwj274

39. Manica M, Bellis AD, Guzzetta G, et al. Intrinsic generation time of the SARS-CoV-2 Omicron variant: An observational study of household transmission. Lancet Reg Health – Eur. 2022;19. doi:10.1016/j.lanepe.2022.100446

40. Ahmadi J, Basiri E, Kundu D. Confidence and prediction intervals based on interpolated records. J Nonparametric Stat. 2017;29(1):1–21. doi:10.1080/10485252.2016.1239826

41. Beutner E, Cramer E. Using linear interpolation to reduce the order of the coverage error of nonparametric prediction intervals based on right-censored data. J Multivar Anal. 2014;129:95–109. doi:10.1016/j.jmva.2014.04.007

42. Kuhn M. caret: Classification and Regression Training. Published online 2022. https://CRAN.R-project.org/package=caret

43. Hegazy N, Cowan A, D’Aoust PM, et al. Understanding the dynamic relation between wastewater SARS-CoV-2 signal and clinical metrics throughout the pandemic. Sci Total Environ. 2022;853:158458. doi:10.1016/j.scitotenv.2022.158458

44. Duvallet C, Wu F, McElroy KA, et al. Nationwide Trends in COVID-19 Cases and SARS-CoV-2 RNA Wastewater Concentrations in the United States. ACS EST Water. 2022;2(11):1899–1909. doi:10.1021/acsestwater.1c00434

45. Li L, Mazurowski L, Dewan A, et al. Longitudinal monitoring of SARS-CoV-2 in wastewater using viral genetic markers and the estimation of unconfirmed COVID-19 cases. Sci Total Environ. 2022;817:152958. doi:10.1016/j.scitotenv.2022.152958

46. Morvan M, Jacomo AL, Souque C, et al. An analysis of 45 large-scale wastewater sites in England to estimate SARS-CoV-2 community prevalence. Nat Commun. 2022;13(1):4313. doi:10.1038/s41467-022-31753-y

47. Hoffmann T, Alsing J. Faecal Shedding Models for SARS-CoV-2 RNA amongst Hospitalised Patients and Implications for Wastewater-Based Epidemiology. Infectious Diseases (except HIV/AIDS); 2021. doi:10.1101/2021.03.16.21253603

48. Crank K, Chen W, Bivins A, Lowry S, Bibby K. Contribution of SARS-CoV-2 RNA shedding routes to RNA loads in wastewater. Sci Total Environ. 2022;806:150376. doi:10.1016/j.scitotenv.2021.150376

49. Arts PJ, Kelly JD, Midgley CM, et al. Longitudinal and quantitative fecal shedding dynamics of SARS-CoV-2, pepper mild mottle virus, and crAssphage. mSphere. 2023;8(4):e00132–23. doi:10.1128/msphere.00132-23

50. Topol A. High Throughput Pre-Analytical Processing of Wastewater Settled Solids for SARS-CoV-2 RNA Analyses V2.; 2021. doi:10.17504/protocols.io.b2kmqcu6

51. Sciences) AT (Verily L, Sciences) BW (Verily L, Michigan) KW (Univ, Boehm AB, marlene.wolfe. High Throughput SARS-COV-2, PMMOV, and BCoV quantification in settled solids using digital RT-PCR. Published online April 21, 2021. Accessed November 22, 2023. https://www.protocols.io/view/high-throughput-sars-cov-2-pmmov-and-bcov-quantifi-btywnpxe

52. Boehm AB, Wolfe MK, White B, Hughes B, Duong D. Divergence of wastewater SARS-CoV-2 and reported laboratory-confirmed COVID-19 incident case data coincident with wide-spread availability of at-home COVID-19 antigen tests. PeerJ. 2023;11:e15631. doi:10.7717/peerj.15631

