## Supplemental Materials for "“Real-time county-aggregated wastewater-based estimates for SARS-CoV-2 effective reproduction numbers”"

*Supplemental Text*

*Method 1: Laboratorial Processing Details*

**Sample collection.** Fifty ml of settled solids were collected in a sterile container daily to three times a week for 6-12 months at 14 POTWs (see Table 1 for precise dates and frequency of sample collection). Additional details of the sample collection can be found in Wolfe et al.^25^ and Kadonsky et al.^27^ Samples were transported to the laboratory at 4°C and processed immediately on receipt at the lab.

**Pre-analytical processing and RNA extraction.** Detailed methods for pre-analytical processing and RNA extraction have been published in other peer-reviewed papers,^13,26^ as well as on protocols.io.^50,51^ In short, solids were dewatered using centrifugation. One aliquot of solids was then used to determine the dry weight and another aliquot of solids was added to DNA/RNA shield (containing spiked in bovine coronavirus, BCoV, as a process control) at a concentration of 75 mg/ml. This concentration was chosen to minimize inhibition of downstream analytical methods.^17,26^ The mixture was homogenized and centrifuged, and nucleic acids were then extracted and purified from the supernatant using a commercial kit. The nucleic acid extract was then processed through an inhibitor removal kit. The resulting RNA was immediately analyzed without storage.

**Analytical measurements.** Undiluted RNA was used as template in a RT-PCR reaction containing previously described primers and probes for a target located in the N gene of SARS-CoV-2,^13,17^ This region has been conserved across all SARS-CoV-2 variants to date.^52^ The RNA was diluted at 1:100 and used as template in a duplex RT-PCR reaction containing previously published primers and probes for pepper mild mottle virus (PMMoV) and BCoV.^13^ The methods used to measure the N gene using digital droplet RT-PCR, including thorough descriptions of the negative and positive controls for both the extraction and PCR, BCoV recovery, QA/QC elements, thresholding methods, and relevant EMMI guideline reporting, have been described in detail in the data descriptor by Boehm et al.^26^ This data descriptor contains a subset of data presented in this paper (including data from Gilroy, Sunnyvale, Palo Alto, San Jose, Silicon Valley, San Francisco Oceanside, San Francisco Southeast, Sacramento, Modesto (Method 1), Merced (Method 1), and Davis collected through 12/31/22). One aspect of the methods differed from those described in the data descriptor for samples collected after 12/31/22; the N gene assay was multiplexed with different assays as the public health needs for wastewater monitoring changed for this prospective monitoring effort. Between January 23, 2023 and March 13 2023, for samples collected from Gilroy/Morgan Hill, Sunnyvale, Palo Alto, San Jose, Silicon Valley, San Francisco Oceanside, San Francisco Southeast, and Sacramento, the N gene assay was multiplexed with an assay for adjacent single nucleotide polymorphisms (SNPs) characteristic of SARS-CoV-2 XBB* in lieu of the SARS-CoV-2 S gene as presented in the data descriptor. For these same sites, between March 13, 2023 and the end of the study, the N gene assay was multiplexed with assays for XBB*, RSV, influenza A, influenza B, and norovirus GII in conjunction with a 6-color droplet reader (QX600, Bio-rad). For the remaining sites, between February 3, 2023 and the end of the study, the N gene assay was multiplex with the XBB* assay. All results are reported as concentrations of the N gene in units of copies per gram dry weight.

*Method 2: Laboratory Processing Details*

**Sample collection.** For Modesto, 250 mL of settled solids were collected in 250-mL HDPE bottles from the primary clarifier sludge outlet four times per week. For Esparto, Woodland, and Turlock, approximately one liter of composite influent was collected using autosamplers four times per week. For Winters, raw untreated wastewater was collected from a pump station using an autosampler four times per week. Sample collection and autosampler details are described in Table S3.

**Pre-analytical processing and RNA extraction.** For Modesto, settled solids were homogenized by inversion. For Esparto, Woodland, Turlock, and Winters, solids were obtained from liquid influent or raw wastewater samples by letting samples settle for a minimum of 20 minutes in a glass beaker. Supernatant was then decanted to retain settled solids. For samples from all Method 2 sites, 50 mL of homogenized sample were dewatered by centrifugation at 24,000 x g for 30 minutes at 4^o^C. Before July 29, 2022, four or five aliquots of 75 mg of dewatered solids were transferred to 50 mL conical tubes, weighed, and diluted in a solution of DNA/RNA shield (containing 1.5 uL of spiked-in BCoV) to achieve a final concentration of 1 mL DNA/RNA shield per 75 mg of dewatered solids. After July 29, 2022, a single aliquot of 750 mg of dewatered solids was diluted in a solution of DNA/RNA shield and BCoV to achieve the same final ratio of 1 mL DNA/RNA shield per 75 mg dewatered solids. Samples were homogenized, and RNA was extracted from 300 uL of homogenized samples using the MagMAX Microbiome Ultra Nucleic Acid Isolation Kit, following manufacturer protocols, using a King Fisher Flex for extractions. Final elution volume was 100 uL and RNA extracts were treated for inhibitor removal using the Zymo OneStep-96 PCR Inhibitor Removal Kit. Silicone plates were prepared by centrifuging plates at 2576 x g for ten minutes, adding samples to plates, and centrifuging plates at 2576 x g for six minutes. RNA extracts were then stored on ice for same-day analysis or stored at -80^o^C for long-term storage.

**Analytical Measurements.** Nucleic acid extracts were used as a template in digital droplet RT-PCR assays. The same primers and probes described for Method 1 for the N gene of SARS-CoV-2, PMMoV, and BCoV were used for Method 2. The N gene was quantified in a triplex assay, alongside the S-gene of SARS-CoV-2 and a SARS-CoV-2 variant-specific mutation target. BCoV and PMMoV were quantified using a duplex assay. Digital droplet RT-PCR was performed on 20 uL samples, aliquoted from 22 uL of total reaction mix, consisting of 5.5 uL of template, 5.5 uL ddPCR One-Step RT-ddPCR Advanced Kit for Probes (catalog no. 1864021; Bio-Rad, CA), 2.2 uL reverse transcriptase, and 1.1 uL of 300 mM dithiothreitol (DTT), and primers and probes at a final concentration of 900 nM and 250 nM, respectively. For triplex assays, 3.3 uL of primer probe mix and 4.4 uL of nuclease-free water were added. For duplex assays, 2.2 uL of primer probe mix and 5.5 uL of nuclease-free water were added. PCR was performed with the following protocol: reverse transcription at 50°C for 60 minutes, enzyme activation at 95°C for 10 minutes, followed by 40 two-step cycles of denaturation at 94°C for 30 seconds and anneal/extension at 58°C (for SARS-CoV-2, PMMoV, and BCoV targets) for 1 minute. This was followed by enzyme deactivation at 98°C for 10 minutes, droplet stabilization at 4°C for 30 minutes, and indefinite hold at 4°C. Droplets were analyzed using the QX200 droplet reader (Bio-Rad). Before May 29, 2022, each sample was run in four replicate wells and from May 30, 2022 onward, each sample was run in five replicate wells. Positive extraction controls and negative extraction controls were each run in one well on each plate. PCR-positive controls and no-template controls (NTC) (negative PCR controls) were each run in one well on each plate. For samples, results from replicate wells were merged for analysis. Thresholding was done using QX Manager Software Regulatory Edition Version 1.2 (Bio-Rad). A minimum of 10,000 partitions were required per well. For a sample to be recorded as positive, at least three positive droplets across all replicates were required.

Multiple controls were included on each plate. Inhibition control consisted of 500,000 copies of BCoV per 75 mg dewatered solids spiked prior to extraction of dewatered solids. A minimum 10% recovery of spiked BCoV was required for sample results to be included. Pepper mild mottle virus (PMMoV) served as a process control. The positive extraction control (PEC) consisted of SARS-CoV-2 genomic RNA (gRNA) (ATCC VR-1986D™ and PolyA: Roche 10108626001) extracted in place of dewatered solids. The negative extraction control (NEC) was prepared by extracting RNA from nuclease free water in place of dewatered solids to identify contamination from extraction processes. Positive controls (PC) for ddPCR consisted of SARS-CoV-2 for the N-gene assay, and BCoV / PMMoV gblock (Integrated DNA Technologies, IDT) for the BCoV and PMMoV assays. PC stock solutions were prepared at a concentration of 50 copies/uL for each target. The no-template control (NTC) consisted of nuclease free water used in place of sample extract to identify contamination during ddPCR plate preparation and analysis.

The concentration of N gene and PMMoV targets in each sample was reported in copies per gram dry weight (cp/g dw), determined as X*B/A*C/Z*S where X is the measured number of copies per ddPCR reaction, B is the total reaction volume, A is the volume of template in the reaction, C is the extract elution volume from the sample extraction kit, Z is the mass of dewatered solids added to each extraction well, and S is the percent solids of the dewatered solids (measured separately for each sample).

*Supplemental Tables*

**Table S1. Overview of minor changes in laboratory methodology for each site during the analysis period.**

| WW Treatment Plants | Dates and method | Dates and method | Dates and method | Dates and method | Dates and method |
| --- | --- | --- | --- | --- | --- |
| San Francisco Southeast, San Francisco Oceanside, Sacramento, Palo Alto, San Jose, Sunnyvale, Silicon Valley, Gilroy/Morgan Hill | 5/1//22-3/12/23    Method 1  Multiplexed as described in data descriptor of Boehm et al.^26^ with 10 replicates per sample | 3/13/22- 5/1/23    Method 1  Multiplexed with assays for  XBB*, RSV, IAV, IBV, HuNoV using the QX600 6 color instrument from Biorad with 10 replicates per sample |  |  |  |
| Esparto, Woodland, and Winters, Turlock | 5/1/2022 – 5/29/2022  Method 2  Triplexed as described in Kadonsky et al.^27^ with 4 replicates per sample. | 5/30/2022 – 11/30/2022  Method 2  Triplexed as described in Kadonsky et al.^27^ with 5 replicates per sample. | 12/2/22-2/6/23    Method 1  Multiplexed with IAV and S with 6 replicates per sample | 2/7/23 - 5/1/23    Method 1  Multiplexed with IAV and  XBB* with 6 replicates per sample |  |
| Davis | 4/20/22-12/31/23    Method 1  Multiplexed as described in in data descriptor of Boehm et al.^26^ with 10 replicates per sample | 1/1/23-2/6/23    Method 1  Multiplexed with IAV and S with 6 replicates per sample | 2/7/23 - 5/1/23     Method 1  Multiplexed with IAV and  XBB* with 6 replicates per sample |  |  |
| Modesto | 5/1/2022 – 5/29/2022  Method 2  Triplexed as described in Kadonsky et al.^27^ with 4 replicates per sample. | 5/30/2022 – 11/30/2022  Method 2  Triplexed as described in Kadonsky et al.^27^ with 5 replicates per sample. | 11/30/22-2/6/23   Method 1  Multiplexed with IAV and S with 6 replicates per sample | 2/7/23 - 5/1/23     Method 1  Multiplexed with IAV and  XBB* with 6 replicates per sample | 2/7/23 - 5/1/23     Method 1  Multiplexed with IAV and  XBB* with 6 replicates per sample |

Note: Description of how details of the analytical measurement of the N gene in SARS-CoV-2 changed during the study period. The changes involve (1) the processing laboratory (defined as either Method 1 or Method 2), (2) the number of replicates from each sample from which nucleic-acids are extracted and subsequently run in individual digital droplet RT-PCR wells, and/or (3) how the N gene assay was multiplexed with other assays. The table provides the name of the sewershed locations in the first column, and then each subsequent column shows the information the progression of method changes from the beginning of the study period to the end. S is the assay targeting the S gene in SARS-CoV-2. IAV is the assay targeting a gene in the influenza A genome. RSV is the assay targeting a gene in the RSV genome. IBV is the assay targeting influenza B. HuNoV is the assay targeting a gene in human norovirus GII. XBB* is the assay targeting 5 adjacent single nucleotide polymorphisms in the XBB* sublineages of SARS-CoV-2.

**Table S2. Overview of attributes for** *R_cc_* **models included in the CalCAT ensemble at the time of writing.**

| *R_cc_* model attributes | Details |
| --- | --- |
| *R_e_* model type | Mechanistic compartmental models, Statistical models, Bayesian estimation |
| Input data streams | Case count, Testing rate, Test positivity, Mortality, Hospitalization, ICU census, Wastewater surveillance |
| *R_e_* nowcast estimation frequency | Daily, Weekly |
| Geographic resolution | County, Region, State |

**Table S3. Wastewater sample collection settings for Method 2.**

| Location | Sample type | Composite sample program |
| --- | --- | --- |
| Esparto | Influent | Time composite: Collected ~10mL every 30 min for 24 hours for weekday samples, and ~10mL every 60 min for 48 hours for weekend samples |
| Turlock | Influent | Flow Composite: Collected ~180 mL for every 415,000 gallons of flow for approximately 24 samples per 24 hour period |
| Modesto | Primary clarifier settled solids | Grab |
| Winters East St. Pump Station | Raw wastewater | Time composite: Collected ~10 mL every 30 min for 24 hours for weekday samples, and ~10mL every 60 min for 48 hours for weekend samples |
| Woodland | Influent | Flow Composite: Collected 100 mL for every 600,000 gallons of flow for approximately 96 samples per 24 hour period |

*Supplemental Figures*

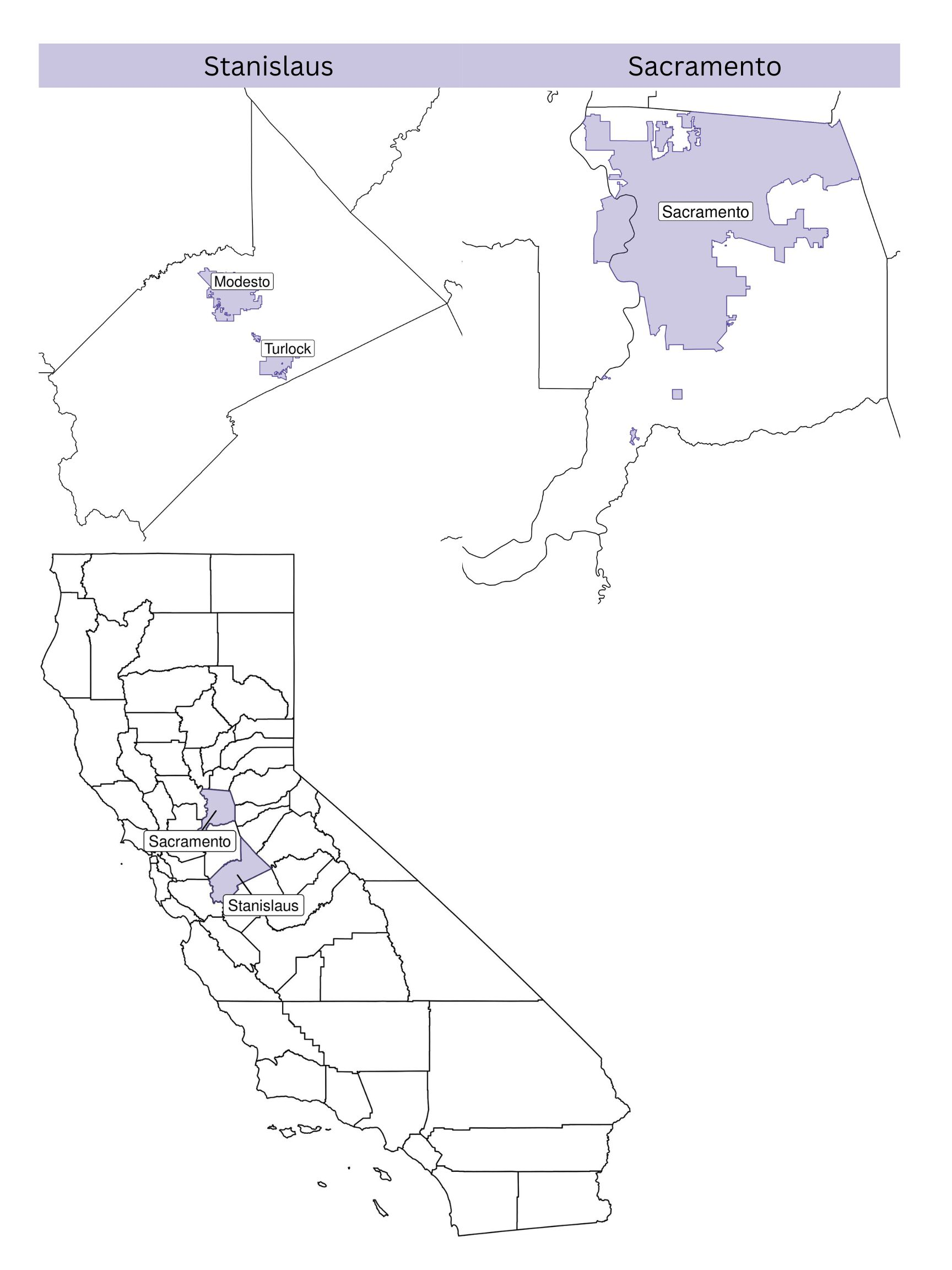

**Figure S1. Map of Sacramento and Stanislaus counties in California and their respective sewersheds that were included in this analysis.**

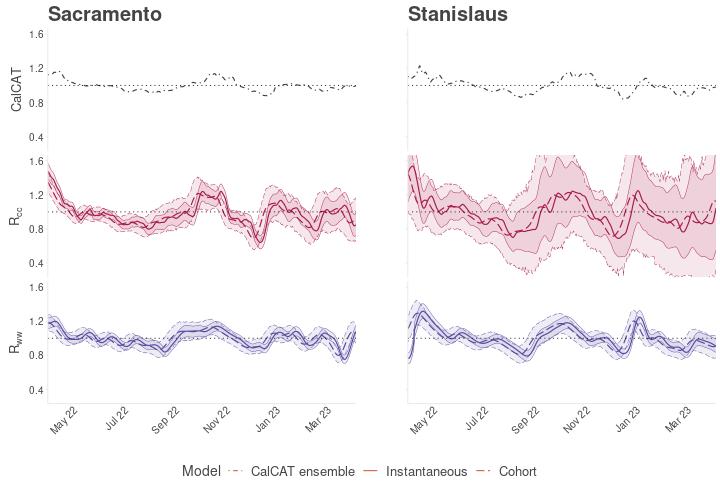

**Figure S2. Time series of county-aggregated, sewershed-restricted *R_e_* for Sacramento and Stanislaus between May 2022 to May 2023**. Three county-level *R_e_* time series are compared: (top, black line) the CalCAT ensemble – a publicly available ensemble of county-wide instantaneous and cohort *R_cc_* estimates; (middle, pink lines) county-aggregated, sewershed-restricted *R_cc_*; and (bottom, blue lines) *R_ww_*. Both *R_ww_* and sewershed-restricted *R_cc_* were calculated using the *R_e_* estimation pipeline piloted in this study. Solid pink or blue lines indicate instantaneous *R_e_*, while dashed pink or blue lines indicate cohort *R_e_*. 95% confidence intervals for each *R_e_* type (instantaneous or cohort) are depicted. Abbreviations: *R_ww_*, sewershed-restricted, wastewater-based effective reproduction number; *R_cc_*, sewershed-restricted case-based effective reproduction number.

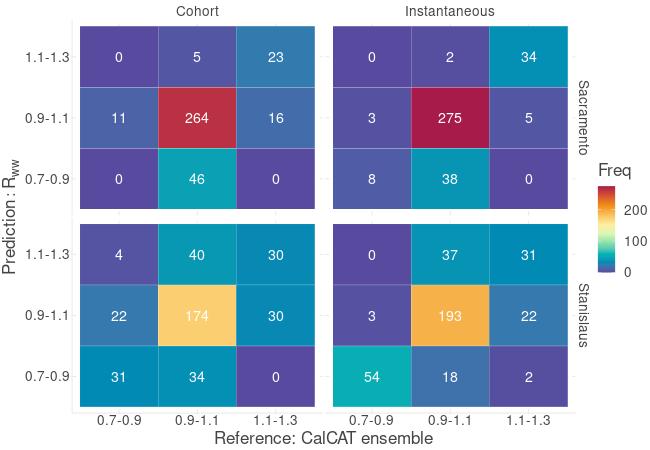

**Figure S3. Confusion matrix depicting the frequency of agreement between** *R_ww_* **and the CalCAT ensemble estimates for Sacramento and Stanislaus counties.** Based on magnitude, *R_e_* values were classified into transmission strength categories (<0.7-0.9, 0.9-1.1, 1.1-1.3, >1.3, which represent a strong decrease, decrease, stability, increase, and strong increase in *R_e_*, respectively). Frequency of agreement between *R_ww_* and the CalCAT ensemble (i.e., instances when predicted *R_ww_* values and reference CalCAT ensemble values belong to the same *R_e_* category) are visualized by the confusion matrix. The right column illustrates results for cohort *R_ww_*, and the left colum illustrates instantaneous *R_ww_* results. Each row represents a single county. The counter diagonals (top right to bottom left) of each matrix represents true positives. Off-diagonal values indicate instances of disagreement between *R_ww_* model predictions and the CalCAT ensemble. Two *R_e_* categories (*R_e_* < 0.7, *R_e_* >1.3) with no *R_e_* values during the study period were excluded. Abbreviations: *R_ww_*, sewershed-restricted, wastewater-based effective reproduction number; Instantaneous, instantaneous *R_ww_*; Cohort, cohort *R_ww_*.

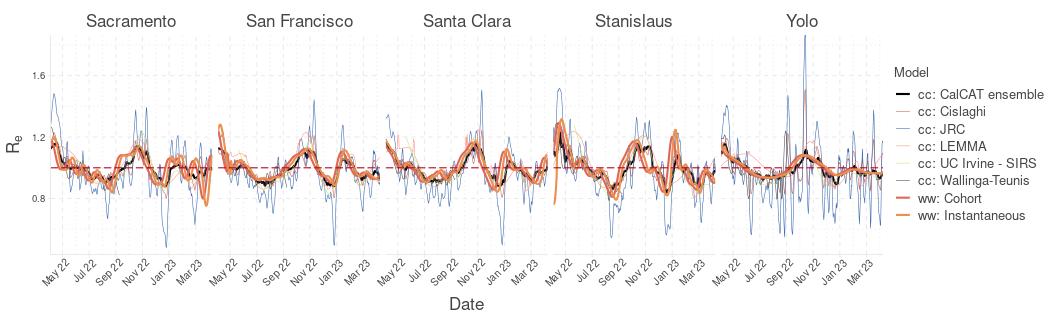

**Figure S4. Comparative time series of** *R_ww_* **versus individual, constituent models of the CalCAT** *R_cc_* **ensemble.**

Abbreviations: cc, case-based effective reproduction number (*R_cc_*); ww, wastewater-based effective reproduction number (*R_ww_*).

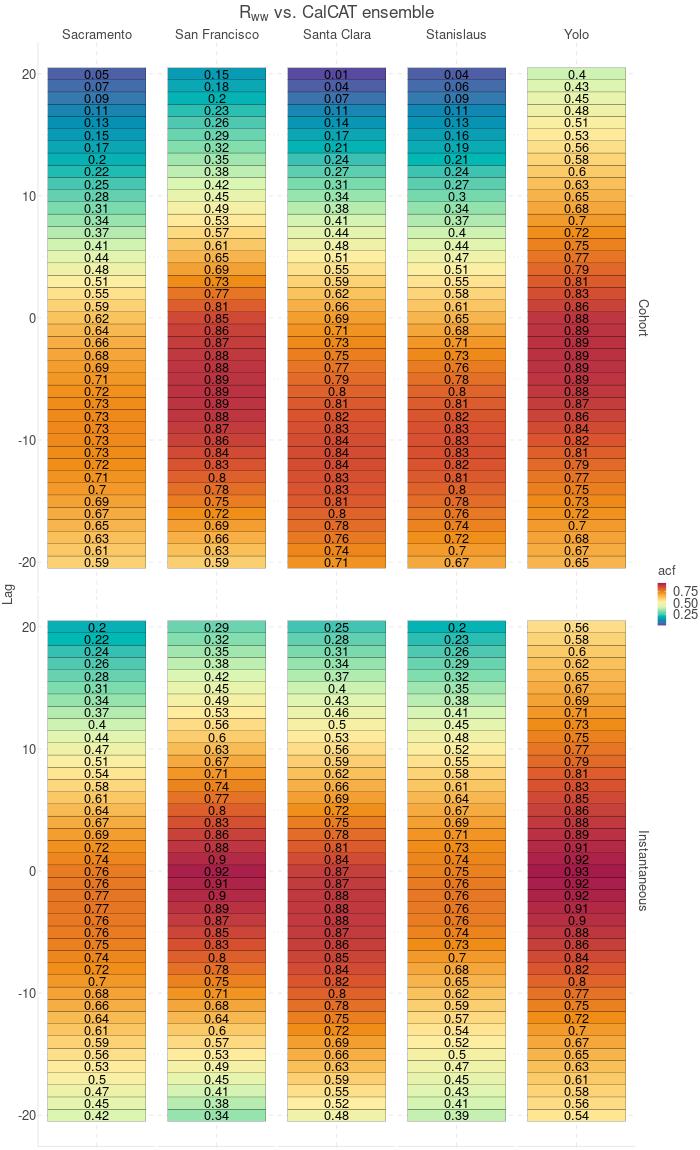

**Figure S5. Heatmap of cross correlations comparing time series of** *R_ww_* **and CalCAT ensemble estimates.**

Using cross-correlation analysis with a maximum lag of 20 days, we investigated temporal alignment of *R_ww_* (cohort or instantaneous) and CalCAT ensemble estimates. A heatmap visualizing cross correlation coefficients for each lag value is shown. Bluer shades represent lower magnitudes of the cross-correlation coefficient, while redder shades represent higher magnitudes of the cross-correlation coefficient. The y-axis corresponds to the range of tested lag values (-20 to 20). Each column corresponds to one of the five studied counties. Negative lag values indicate *R_ww_* temporally precedes the ensemble**;** positive lag values indicate the ensemble temporally precedes *R_ww_*; lag values of zero indicate no temporal shift of *R_ww_* with respect to the ensemble. Abbreviations: *R_ww_*, sewershed-restricted, wastewater-based effective reproduction number; Instantaneous, instantaneous *R_ww_*; Cohort, cohort *R_ww_*; CC, cross-correlation coefficient

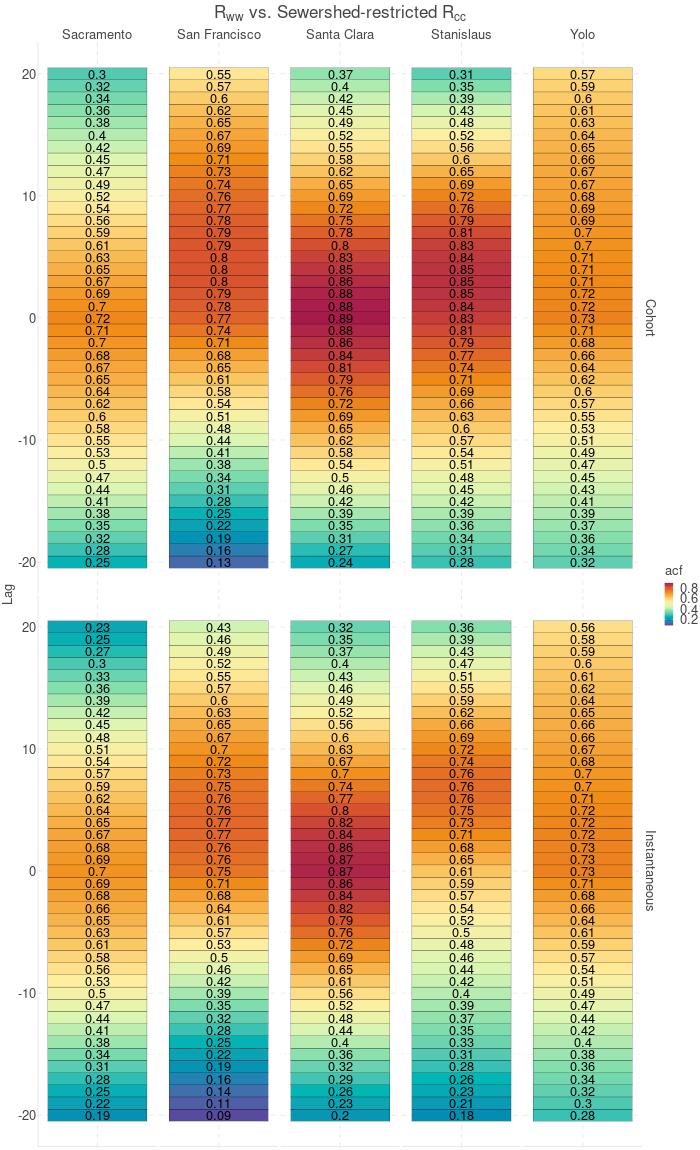

**Figure S6. Heatmap of cross correlations comparing time series of** *R_ww_* **and sewershed-restricted** *R_cc_***.**

Using cross-correlation analysis with a maximum lag of 20 days, we investigated temporal alignment of *R_ww_* (cohort or instantaneous) and sewershed-restricted *R_cc_*. A heatmap visualizing cross correlation coefficients for each lag value is shown. Bluer shades represent lower magnitudes of the cross-correlation coefficient, while redder shades represent higher magnitudes of the cross-correlation coefficient. The y-axis corresponds to the range of tested lag values (-20 to 20). Each column corresponds to one of the five studied counties. Negative lag values indicate *R_ww_* temporally precedes *R_cc_***;** positive lag values indicate the ensemble temporally precedes *R_cc_*; lag values of zero indicate no temporal shift of *R_ww_* with respect to the *R_cc_*. Abbreviations: *R_ww_*, sewershed-restricted, wastewater-based effective reproduction number; sewershed-restricted *R_cc_*, sewershed-restricted case-based effective reproduction number; Instantaneous, instantaneous *R_ww_*; Cohort, cohort *R_ww_*; CC, cross-correlation coefficient

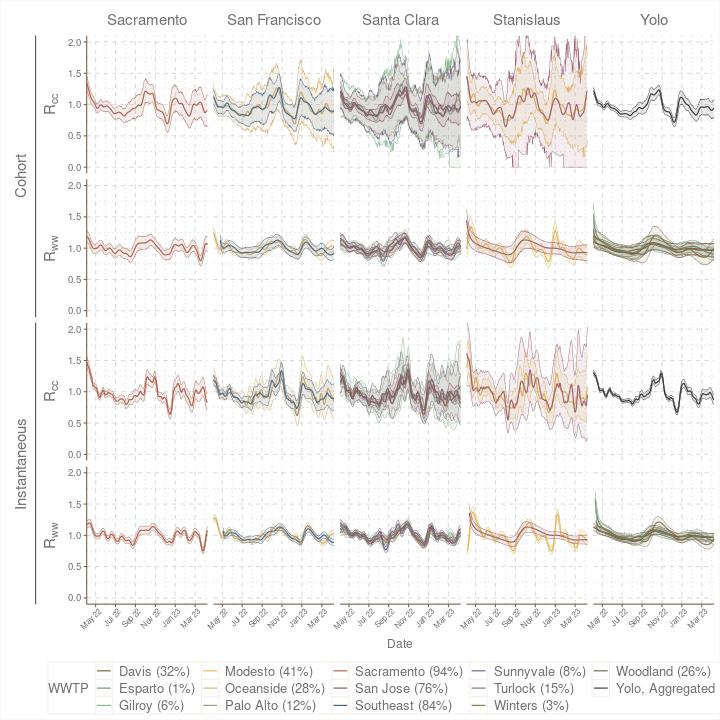

**Figure S7. Individual sewershed-level** *R_cc_* **and** *R_ww_****­* estimates stratified by county (columns) and estimation method (rows).** These individual sewershed-level *R_e_*, which are ultimately population-weighted and aggregated to yield county-level *R_e_* . Percentages in legend indicate proportion of total county population surveilled by the corresponding wastewater treatment plant. Abbreviations: WWTP, wastewater treatment plant; *R_ww_*, wastewater-based effective reproduction number; *R_ww_*, sewershed-restricted, wastewater-based effective reproduction number; *R_cc_*, sewershed-restricted case-based effective reproduction number; *R_e_ ,* effective reproduction number; Instantaneous, instantaneous *R_e_*; Cohort, cohort *R_e_*.
